# CSF turnover reshapes biomarker interpretation in neurodegeneration studies

**DOI:** 10.64898/2026.02.02.26345363

**Authors:** Pablo García-González, Raquel Puerta, Jonas Dehairs, Chengran Yang, Ciyang Wang, Jigyasha Timsina, Itziar de Rojas, Claudia Olivé, Alejandro Valenzuela, Paula Bayón-Buján, Marta Rovira, Laura Montrreal, Maria Capdevila, Álvaro Muñoz-Morales, Berta Calm, Sergi Valero, Montse Alegret, Marta Marquié, The Global Neurodegeneration Proteomics Consortium (GNPC), John C. Morris, Suzanne E. Schindler, David M. Holtzman, Pilar Sanz, Lluís Tárraga, Asif Khan, Maria E. Sáez, Bart Smets, Adelina Orellana, Xavier Montalbán, Mercè Boada, Amanda Cano, Menghan Liu, Muhammad Ali, Carlos Cruchaga, Johannes V. Swinnen, Victoria Fernández, Alfredo Cabrera-Socorro, Agustín Ruiz

## Abstract

Cerebrospinal fluid (CSF) biomarkers are central to Alzheimer’s disease (AD) diagnosis and research. However, CSF composition is shaped not only by neurodegeneration, but also by underlying physiological and pathological processes that remain poorly characterized. By integrating multi-omics data from the deeply characterized memory-clinic ACE CSF cohort (N=1,372), the Global Neurodegeneration Proteomics Consortium (N=1,863), and publicly available quantitative trait *loci* data, we reveal that 73.2-85.9% of the molecular variance in CSF omics data is driven by two main factors: one reflecting CSF turnover rate, and another representing blood-brain barrier (BBB) integrity. CSF turnover mainly determines brain-derived molecules, while BBB damage leads to increased blood-derived protein abundance. CSF turnover/clearance severely impacted core AD biomarker levels, affecting the classification of subjects in the A/T framework. Adjusting biomarker levels for OPCML, a novel reference marker, improved biomarker-based prediction of AD progression and removed confounded associations, revealing a proteomic signature of sporadic AD pathology that closely resembles that of autosomal dominant AD. Finally, using the ACE CSF cohort as discovery (N=1,221) and Knight ADRC as replication (N=1,073), we report a curated AD signature comprising 446 unique proteins. Our findings identify CSF dynamics as a major source of molecular variation, reshaping the interpretation of CSF biomarkers.

## Main

Cerebrospinal fluid (CSF) surrounds and fills the brain and spinal cord, providing mechanical protection, nutrition, chemical homeostasis and waste removal to the central nervous system (CNS)^1,2^. In the absence of routine brain biopsies, CSF can be collected non-invasively via lumbar puncture, representing the closest approximation to CNS tissue molecular pathobiology and enabling large-scale studies in living subjects. In the context of Alzheimer’s Disease (AD), CSF levels of amyloid beta 42 (Aβ42) and phosphorylated tau (p-tau), commonly referred to as “core AD biomarkers”, are widely used in clinical and research settings to report the presence of AD pathology in the brain^3–5^.

Understanding CSF physiology is crucial for interpreting biomarker variation in this biofluid correctly^1,2,6^. CSF is actively secreted by the choroid plexus (CP), a heavily vascularized structure located within the brain ventricles, and it is primarily composed of ions, peptides, proteins and micronutrients^7^. CSF flows through the ventricles into the subarachnoid space, where it is distributed to the rest of the brain or into the lumbar sac^8^. Once in contact with the brain parenchyma, molecular exchange occurs by diffusion between the parenchymal interstitial fluid and the CSF, allowing clearance of molecular products and altering CSF composition. Finally, CSF exits the CNS via the arachnoid granulations into the blood, or via the nasal cribriform plate drainage into the lymph system^9^. Importantly, factors influencing CSF turnover in the brain—including sleep, circadian oscillations, CP hyperplasia, drugs and physiological factors—as well as blood-brain-barrier (BBB) integrity, can dramatically alter CSF composition independently of underlying brain pathology^1,2,6,10^. These factors likely represent latent features of CSF molecular variability, which are rarely accounted for in biomarker studies^11–14^.

Given the growing relevance of CSF biomarkers in neurodegeneration research and clinical practice^5,11–24^, assessment of the physiological and pathological factors that influence CSF composition is crucial to ensure adequate study design and interpretation. Importantly, two prior studies have reported substantial, non-disease-related, inter-individual variability in CSF protein levels ^25,26^, although the underlying causes, and whether it reflects one or multiple latent biological processes still remain unclear.

Here, we used a data-driven approach to dissect the main sources of variance in CSF molecular composition in the deeply characterized ACE CSF cohort (N=1,372; Extended Data 1). Integrative analysis of proteomics (Somascan 7K), lipidomics (Lipometrix), AD core biomarkers (ELISA/Lumipulse), and CSF ancillary measures (e.g. turbidimetry) revealed shared patterns of molecular variability. To identify the biological drivers of these cryptic components, we combined proteomic signatures, genome-wide association analysis, quantitative trait locus (QTL) data, pathway enrichment and gene expression profiling. Proteomic profiles were further validated in four independent cohorts from the Global Neurodegeneration Proteomics Consortium^27^ (GNPC; N=1,863). Adjusting AD core biomarkers with reference proteins capturing these molecular patterns improved disease outcome prediction and reduced statistical inflation in proteomic analyses. Finally, we used adjusted AD biomarker data to define a robust CSF proteomic signature of AD pathology, replicated in the Knight ADRC cohort^28^ (N=1,073).

## Results

### Principal component analysis

A summary of the ACE CSF cohort and data quality control procedures (QC) is available in the methods section (Extended Data 2, Supp. Table 1-3). After QC, we leveraged a lipidomics dataset including 386 lipid species (*N=*1,176), and a proteomics dataset including 2,395 protein measurements (*N=*1,289). The standardized levels of CSF analytes in the lipidomics and proteomics experiments were highly inter-correlated, revealing a pattern of inter-individual variability (Fig. 1). The overall intensity, which we quantified as the mean standardized values (MSV) of all measured species in each sample, was strongly correlated between both omics experiments (*β*=0.61; *p*=8.7·10^-113^, *R*^2^=0.37), indicating this parameter reflected an intrinsic property of the CSF rather than just experimental noise (Fig. 1). To disentangle the cryptic patterns driving this variability, we conducted principal component analysis (PCA) in both omics datasets separately. The first two principal components (PC1 and PC2) combined explained 75% of the lipidome and 74% of the proteome variance respectively (Fig. 2a). Furthermore, the first two PCs were the most relevant factors explaining the MSVs in both omic datasets (Supp. Fig. 1). Additionally, the first (*β*=0.75; *p*=1.7·10^-205^, *R*^2^=0.57) and second (*β=*0.47; *p*=1.5·10^-62^, *R*^2^=0.22) PCs were highly correlated between both omic experiments, while most combinations of other PCs and intensities yielded milder or null correlation coefficients (Fig. 2b). These observations are consistent with the presence of a common latent biological process that concurrently shapes protein and lipid levels in human CSF.

**Figure 1.**
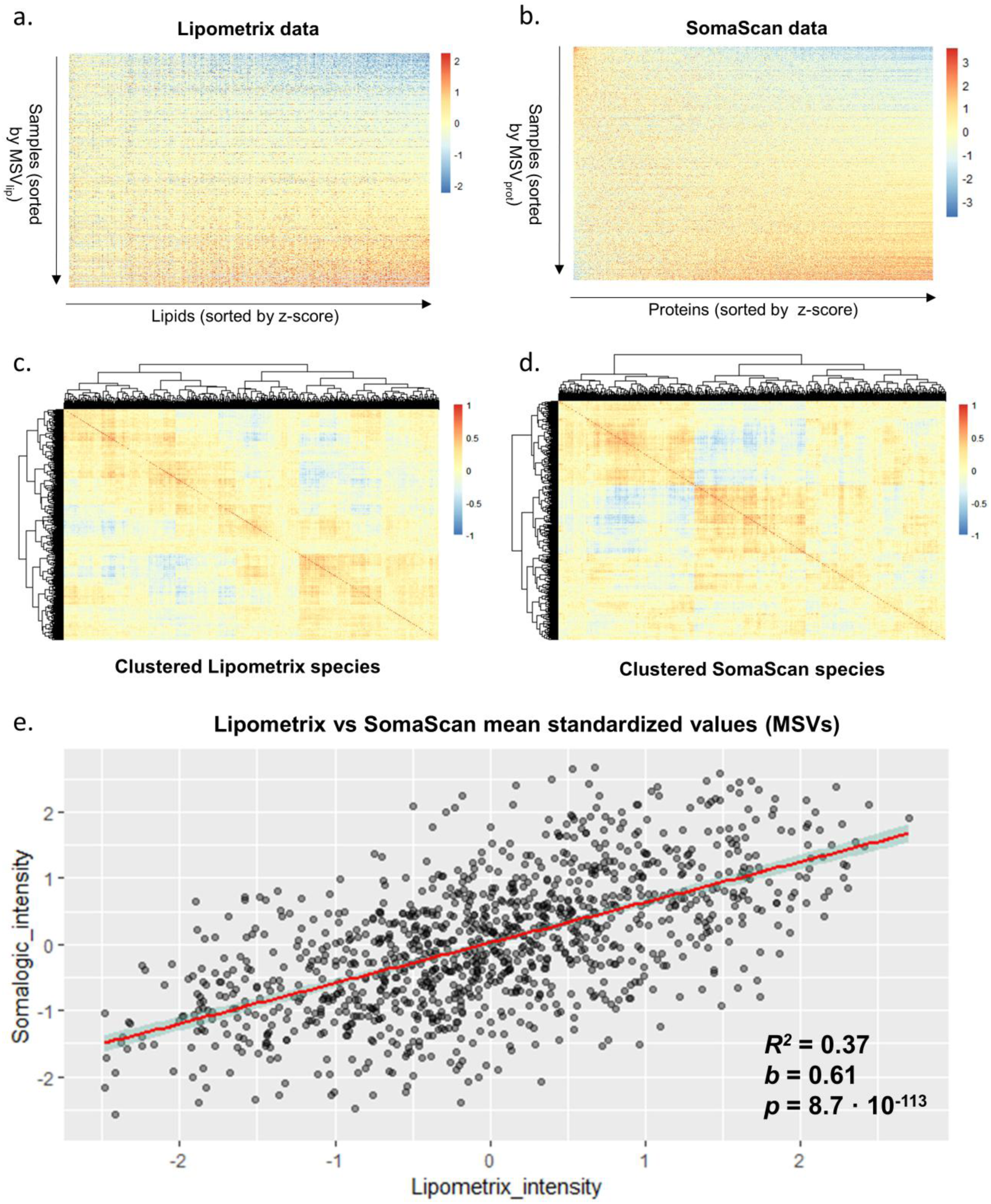
Overview of CSF proteomics and lipidomics data in the ACE CSF cohort. **a-b**. standardized values of all analytes in the lipidomics (N=1,176 samples, 386 species) and proteomics (N=1,289; 2,395 species) datasets, respectively. Rows represent samples, sorted by their mean standardized intensities. Columns represent lipid/protein species, sorted by z-values derived from linear regressions fitting each species to the mean standardized intensity. **c-d.** Hierarchically clustered correlation heatmap of the lipid and protein species in each dataset, respectively. E. Correlation between the Lipometrix and SomaScan MSVs.

**Figure 2.**
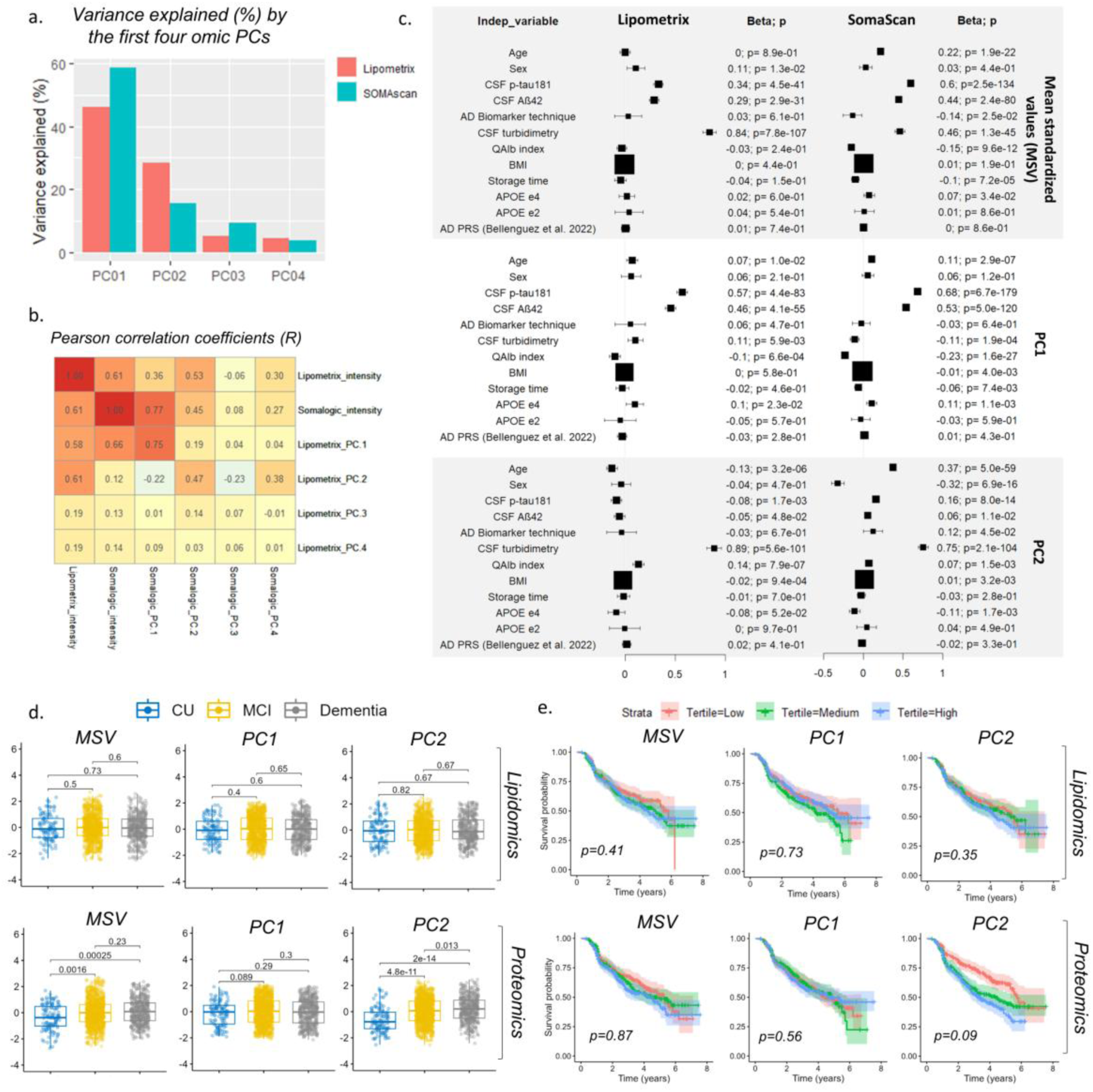
PCA summary and characterization. **a.** Variance explained (%) by the first four proteomics and lipidomics PCs. **b.** Pearson correlation coefficients between the MSVs (intensity) and the first four omic PCs. **c.** Forest plot displaying the associations found for the MSVs and first two PCs with biochemical features (CSF turbidimetry, QAlb index), demographics (age at lumbar puncture, sex), AD biomarkers (Aβ42, p-tau181), genetic features (*APOE* ε4 and ε2 dosage, AD polygenic risk score), and other relevant variables such as the body mass index (BMI) and sample storage time. AD Biomarker Technique employed to determine CSF Aβ42 and p-tau181 levels (ELISA/Lumipulse) was included as a covariate in the models to adjust for potential technical variability. **d.** Distribution of the MSV and first four PCs across the dementia continuum (CU, MCI, Dementia) in the ACE cohort. Brackets display two-sided p-values derived from pairwise Wilcoxon rank-sum tests. **e.** Kaplan-Meier plots showing MCI to AD conversion survival curves of tertile-split individuals (N=632 Lipometrix; N=737 SomaScan). The p-values represent those derived from Cox models adjusted by age at baseline and sex.

### Characterization and clinical relevance of CSF omic PCs

To better understand the mechanisms underlying CSF molecular variance, we ran multivariate linear regression models including biochemical features (CSF turbidimetry, QAlb index), demographics (age, sex), AD biomarkers (CSF Aβ42 and p-tau181), genetic features (*APOE* ε4 and ε2 dosage, AD polygenic risk score), and other relevant variables such as the body mass index (BMI) or sample storage time. Our results indicated a strong positive association of p-tau181 and Aβ42 levels with MSVs and PC1s in both omics datasets (Fig.2c). Conversely, the PC2 showed weaker or null associations with the AD biomarkers and exhibited a strong association with CSF turbidimetry measurements, suggesting it may capture pathophysiological mechanisms related to blood-brain barrier (BBB) damage. The proteomics PC2 was also strongly increased with older age (*p*=5.0·10^-59^) and male sex (*p*=6.9·10^-16^), Finally, sample storage time, blood cell count and paired CSF/serum biochemistry variables showed little to no association with the CSF omic PCs (Fig. 2c, Supp. Fig. 2-3), suggesting that these major PCs were driven by subject pathophysiology, rather than blood contamination of the CSF during lumbar puncture, or protein degradation due to long-term biobanking.

While lipidomics cryptic phenotypes were evenly distributed across clinical groups, the proteomics PC2 increased progressively along the dementia continuum (Fig. 2d), further stressing BBB damage as a potential driver. However, neither MSV, PC1 or PC2 were associated with mild cognitive impairment (MCI) to AD-dementia progression (Fig. 2e). Interestingly, subsequent components of the proteomic variance (PC3 [*HR=*1.36; *pBonf=*1.47·10^-06^], PC7 [*HR=*0.52; *pBonf=*3.13·10^-27^], and PC9 [*HR=*1.27; *pBonf=*1.62·10^-03^]) were strongly associated with disease progression (Supp. Table 4), potentially capturing molecular signatures associated with AD pathogenesis.

### Genetic determinants of CSF multi-omic PCs

To gain insights into the potential biological mechanisms underlying the cryptic phenotypes, we conducted genome-wide association studies (GWAS) of the MSV and the first ten omic PCs (Supp. Table 5). The *GMNC* locus was genome-wide significant (GWS; p<5·10^-08^) for MSVs and PC1 in both omics datasets (Fig. 3). Furthermore, we identified GWS signals in the *FADS1/FADS2 locus* (lipidomics PCs 6 to 8), *PRICKLE1* (lipidomics PC8) and *APOE* (proteomics PC7 and PC9), with SNP tagging the *ε4* allele (rs429358), as the most significant variant in the *locus* (Table 1). *APOE* is a recognized pleiotropic trans-regulator of the proteome both in plasma and CSF^29–31^. Similarly, the *FADS1*/*FADS2 locus* is a well-established pleiotropic regulator of lipid levels in plasma, CSF and brain ^32,33^. A GWS signal was also detected in chromosome 16p12.3, approximately 250 kb downstream of the closest protein-coding gene *NPIPA8*, for proteomics PC9.

**Table 1.** Summary of GWS hits found within the MSVs and the first ten PCs of the lipidomics (N=1,087) and proteomics (N=1,226) datasets. Genomic coordinates correspond to the *GRCh38 build. A1, effect allele; A2, other allele; A1_FREQ, frequency of the effect allele; N, GWAS sample size; Z, z-score; P, p-value*.

| | Phenotype | $\lambda_{GC}$ | Locus | rsid | Cytogenetic band | A1 | A2 | A1_FREQ | Z | P |
| --- | --- | --- | --- | --- | --- | --- | --- | --- | --- | --- |
| Lipometrix | MSV | 1.01 | *GMNC* | rs34596688 | chr3q28 | C | CT | 0.41 | 5.79 | $9.21 \cdot 10^{-9}$ |
| | PC1 | 1.00 | *GMNC* | rs9842055 | chr3q28 | G | T | 0.47 | 5.70 | $1.55 \cdot 10^{-8}$ |
| | PC6 | 1.00 | *FADS1/FADS2* | rs174560 | chr11q12.2 | C | T | 0.26 | 12.46 | $2.23 \cdot 10^{-33}$ |
| | PC7 | 0.99 | *FADS1/FADS2* | rs174549 | chr11q12.2 | A | G | 0.24 | -8.11 | $1.32 \cdot 10^{-15}$ |
| | PC8 | 1.01 | *FADS1/FADS2* | rs5792235 | chr11q12.2 | C | CA | 0.28 | 7.27 | $7.05 \cdot 10^{-13}$ |
| | | | *PRICKLE1* | rs17488421 | chr12q12 | T | A | 0.08 | -5.86 | $6.13 \cdot 10^{-9}$ |
| | PC10 | 1.02 | *FADS1/FADS2* | rs174544 | chr11q12.2 | A | C | 0.24 | -5.60 | $2.67 \cdot 10^{-8}$ |
| SomaScan | MSV | 1.02 | *GMNC* | rs34596688 | chr3q28 | C | CT | 0.41 | 7.03 | $3.48 \cdot 10^{-12}$ |
| | PC1 | 1.02 | *GMNC* | rs59966036 | chr3q28 | T | TG | 0.40 | 5.61 | $2.54 \cdot 10^{-8}$ |
| | PC7 | 1.00 | *APOE* | rs429358 | chr19q13.32 | C | T | 0.20 | -30.67 | $1.44 \cdot 10^{-153}$ |
| | PC9 | 1.01 | *APOE* | rs429358 | chr19q13.32 | C | T | 0.20 | 11.04 | $4.58 \cdot 10^{-27}$ |
| | | | *16p12.3* | rs76649197 | chr16p12.3 | G | T | 0.08 | -5.64 | $2.17 \cdot 10^{-8}$ |

**Figure 3.**
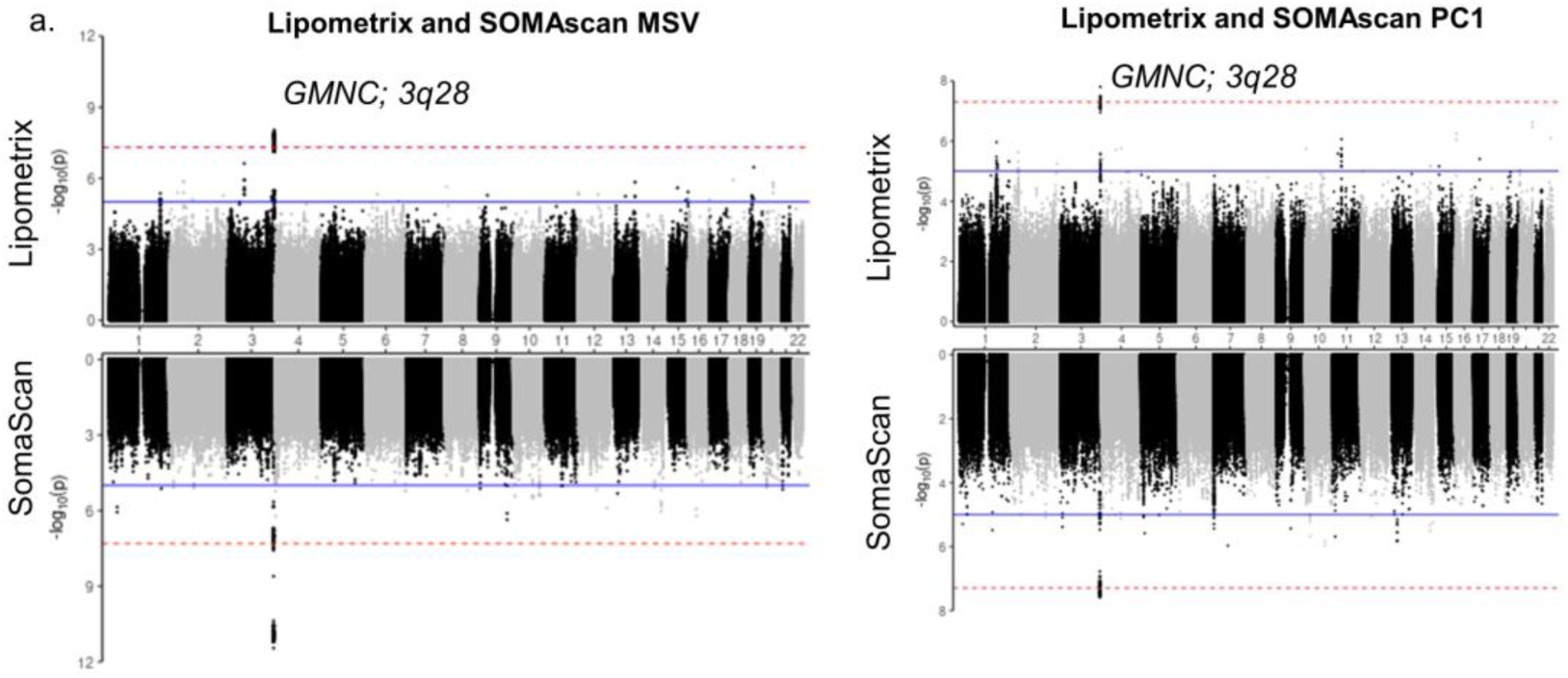
GMNC is a quantitative trait locus for our MSV and PC1 phenotypes in both lipidomics and proteomics data. Miami plots showing GWAS results for MSV and the first PC of the Lipometrix and SomaScan data in the ACE CSF cohort. The *GMNC* locus was genome-wide significant (*p*<5·10^-08^) for all these phenotypes.

*GMNC* is a well-recognized *locus* affecting brain lateral ventricular volume^34^ and CSF p-tau levels^35–38^. In the brain, *GMNC* is highly expressed in the choroid plexus (CP) epithelium and ependymal cells (Supp. Fig. 4)^39^, and functions as a differentiation master regulator of multiciliated cells, the cells driving fluid clearance across epithelia^40^. This finding led to a mechanistic hypothesis: genetic variants enhancing CSF production cause increased CSF turnover through the brain, simultaneously enhancing clearance of brain-derived molecules while promoting ventricular expansion. Indeed, five of the seven ventricular volume increasing alleles (VVIAs) identified in a previous GWAS^34^ (*GMNC, C16orf95, AMZ1/GNA12, TRIOBP* and *MLLT10*) were associated with a decrease in PC1, MSV, p-tau181, CSF turbidimetry and Aβ40 levels (Fig. 4). These results support that genetic predisposition to larger ventricular volumes correlates with lower CSF concentrations (Fig. 4a).

**Figure 4.**
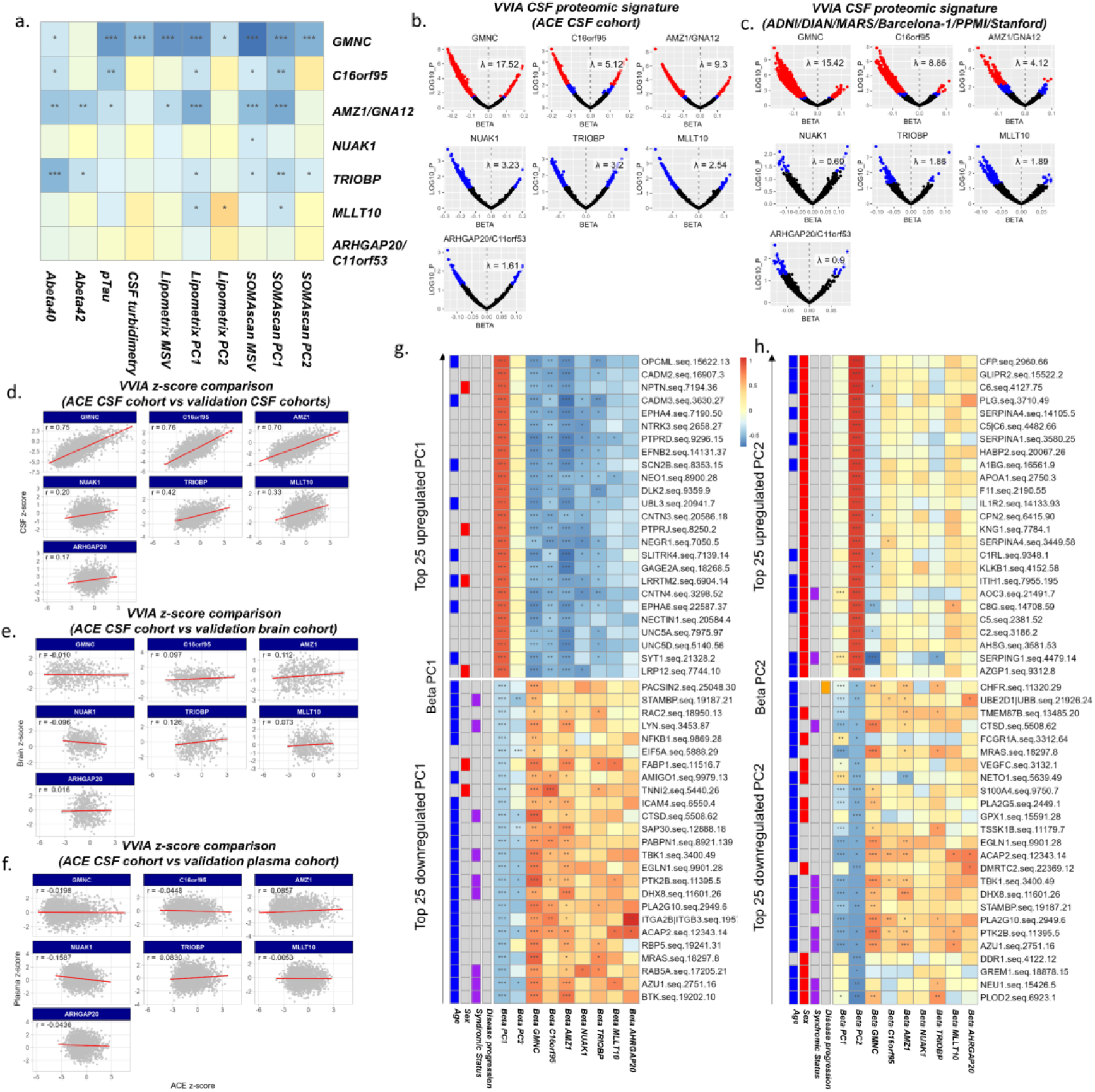
Proteogenomic characterization of CSF multi-omic PC1 and PC2. **a.** z-scores of ventricular volume increasing alleles (VVIAs) across reported *loci*. **b-c.** Volcano plots showing effect sizes (betas) and –log10 transformed p-values for associations between VVIAs and proteins in the CSF Somascan datasets for the ACE CSF cohort (*N=*1,226) (b), and the validation cohorts ADNI/DIAN/MARS/Barcelona-1/PPMI/Stanford (*N=*2,784) (c). **d-f.** Correlation of Somascan z-scores for each VVIA, comparing results between the ACE CSF cohort and validation cohorts in CSF (*N=*2,784), brain (*N=*378), and plasma (*N=*2,338), respectively. r displays the Pearson correlation. **g-h.** Integrated proteogenomic signatures of CSF multi-omic PC1 (g) and PC2 (h) in the ACE CSF cohort. We represented the 25 somamers most positively and negatively correlated with each PC, with annotations indication Bonferroni-significant associations with age, sex, disease group and disease progression. To account for differences in beta ranges across tests, we standardized betas (SD=1) to enhance visual representation. VVIAs were defined as: *GMNC* (rs34113929-G), *C16orf95* (rs9937293-A), *AMZ1/GNA12* (rs798562-T), *NUAK1* (rs12146713-C), *TRIOBP* (rs4820299-C), *MLLT10* (rs35587371-T), and *ARHGAP20/C11orf53* (rs7936534-A). \**p*<0.05, \*\**p*<0.01, \*\*\**p*<0.001.

### VVIAs cause a CSF-specific molecular signature

To further explore our hypothesis, we checked the impact of VVIAs on individual lipid and protein species within our datasets. The resulting signatures were highly inflated and asymmetric, with a striking predominance of species displaying decreased abundance in the presence of VVIAs (Table 2, Fig. 4b, Extended Data 3-4). Interestingly, a distinct protein subset showed the opposite trend (Extended Data 4), suggesting more complex dynamics than a simple dilution effect caused by increased CSF production.

**Table 2:**
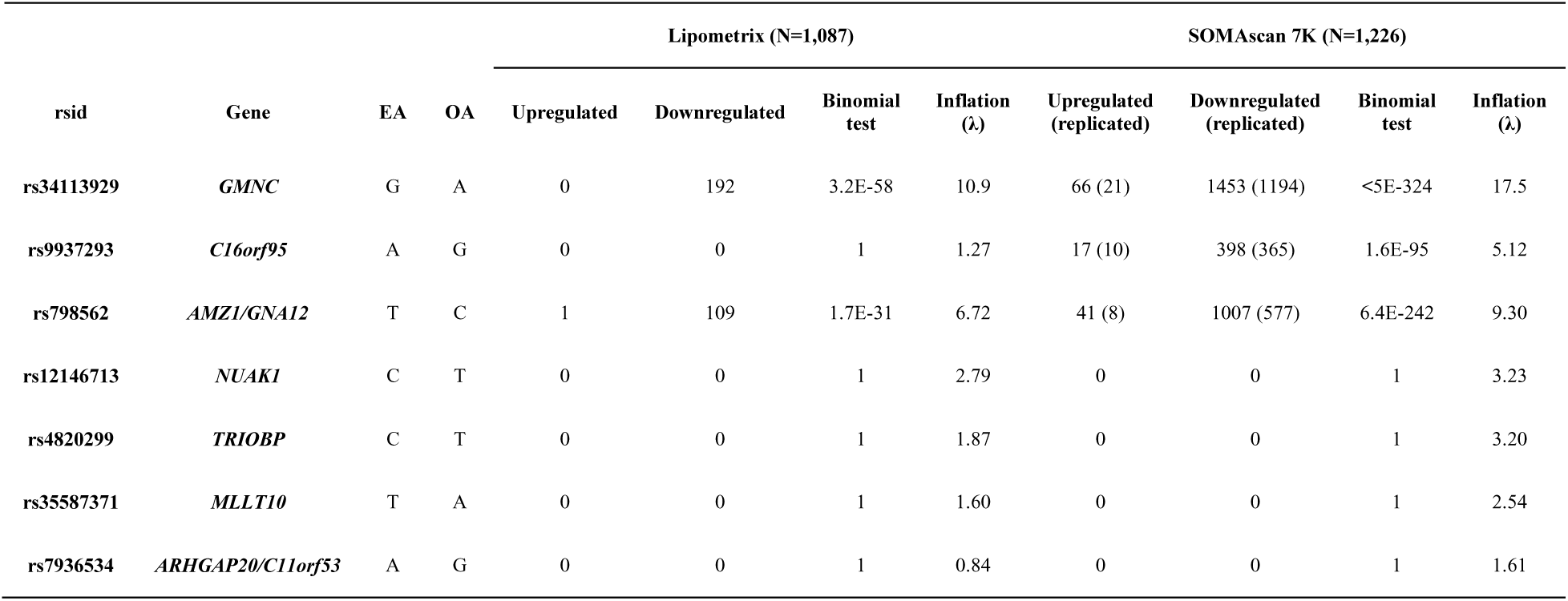
Number of FDR-significant up- and downregulated lipid species and proteins by ventricular volume-increasing alleles in the ACE CSF cohort. , including a binomial test p-value comparing the number of up- versus downregulated FDR-significant analytes, and the lambda inflation values. For Somascan analytes, we also disclose the number of replicated somamers in the QTL data used for validation^30^.

Then, we validated these findings using proteomic (Somascan) and metabolomic (Metabolon) QTL inventories in three different tissues—CSF, brain and plasma (Supp. Table 7-8)^30,33,41,42^. CSF VVIA signatures were again characterized by negatively skewed beta coefficients and strong inflation (Fig. 4c). We replicated a large proportion of the FDR-significant associations found in the ACE CSF cohort (Table 2), with highly correlated z-score distributions across cohorts (Fig. 4d). Furthermore, z-scores were also strongly correlated across different VVIAs, indicating that these independent *loci* exert a similar influence on CSF composition (Supp Fig. 5-6). In contrast, VVIAs had no influence on the brain and plasma multi-omic studies explored (Fig. 4e-f, Supp Fig. 5-6, Supp. Table 7). These findings reinforce the notion that PC1 represents a CSF-specific phenotype, unimpacted by neural or blood tissue pathophysiology.

### PC1 represents CSF turnover rate

To gain further insight, we explored the Somascan dataset (2,395 somamers), integrating multi-omic PC proteomic signatures (see methods) with their corresponding VVIA-somamer associations. Given the large number of significant proteins (Supp. Fig. 7-8), we focused mainly on the top 25 upregulated and downregulated proteins associated with PC1. These proteins showed significant and directionally opposite associations with VVIAs (Fig. 4g). GTEx gene expression profiling revealed that PC1-upregulated proteins were highly expressed in the brain, while, in comparison, PC1-downregulated proteins showed higher expression in whole blood and leukocytes (Extended Data 6). Top PC1-upregulated proteins, including OPCML, CADM2, and NPTN, exhibited the strongest absolute correlations (inferred by squared beta values). Downregulated proteins included BTK, a crucial component of B cell signalling^43^, and AZU1, a well-known antimicrobial protein^44^. Geneset enrichment analysis (GSEA) of the PC1 signature identified upregulated pathways related to synaptic function, neuronal processes and the connectome; while downregulated pathways were related to immune functions and plasma-derived components (Extended Data 7). Integrative GSEA combining the VVIA proteomic signatures for *GMNC, C16orf95*, and *AMZ1/GNA12* revealed similar, but directionally opposite, enrichments, further supporting an inverse relationship between PC1 and ventricular volume (Extended Data 7).

These findings reinforce our previous hypothesis: elevated CSF turnover likely leads to dilution of proteins and metabolic waste generated within the CNS. The smaller subset of the proteome downregulated by PC1 and upregulated by VVIAs (increased abundance due to higher CSF turnover) likely represent proteins actively transported across the blood-CSF barrier by the choroid plexus. These proteins include immune and signalling factors potentially targeted to the CNS. In line with this, leptin and prolactin, two proteins transported from blood to CSF through the CP^1,45^, were among this small group of PC1-downregulated, VVIA-upregulated proteins (Extended Data 8).

### PC2 represents BBB damage

In contrast, the top 25 PC2-upregulated proteins showed minimal regulation by VVIAs (Fig. 4g), suggesting a different underlying biological mechanism. Most of these proteins are synthetized by the liver (Extended Data 6) and included complement cascade proteins (e.g. CFP, C6 and C5|C6 complex), plasma lipoproteins (APOA1), or proteins related to coagulation (PLG, SERPINA1 and F11; Fig. 4g). GSEA highlighted complement activation and blood microparticle signatures among the upregulated pathways in PC2 (Extended Data 7). The downregulated portion of PC2 mostly overlapped with PC1-regulated proteins (Supp. Fig. 8) and exhibited weak statistical significance (Fig. 4g); therefore, we did not pursue further characterization.

Our results suggest that PC2 may represent plasma leakage to the CSF via BBB damage. Protein concentration in the blood is hundreds of times higher than in the CSF^10^, explaining the trend of PC2 toward increased protein abundance. Additionally, the proteomics PC2 was associated with patient history of cerebrovascular accident (p=7.9·10^-04^), cardiopathy (p=2.7·10^-04^), diabetes (p=1.8·10^-08^), hypertension (p=8.3·10^-12^) and renal insufficiency (p=1.4·10^-06^, Supp. Fig. 9), in addition to male sex, older age, and syndromic status (Fig. 2). This pattern resembles the connection between BBB integrity and dementia, potentially mediated through cardiovascular dysfunction. However, we found no associations between PC2 and genetic predisposition to stroke^46^ or hypertension^47^ (Supp. Fig. 10), limiting causal inference. A mild association was present between red blood cell count in the CSF and proteomics PC2 (Supp. Fig. 9), suggesting a modest contribution of traumatic lumbar puncture, although this association was lost when adjusting for CSF total protein concentration (Supp. Fig. 3).

### PC1 and PC2 signatures are consistent across four GNPC cohorts

Analysis of four additional CSF cohorts from the Global Neurodegeneration Proteomics Consortium (GNPC; *N=*1,863)^27^ revealed similar patterns of inter-individual variability, with the first two PCs explaining 73.2% to 85.9% of the total proteomic variance across the different cohorts (Supp. Fig. 11). At the protein level, PC1 and PC2 signatures were also highly consistent, closely reproducing the beta distributions across somamers found in the ACE CSF cohort (Extended Data 9, Supp. Table 8). Top markers also replicated strongly: OPCML (PC1) emerged as the leading marker in two GNPC cohorts (Contributors N and T), with consistent effect sizes across all cohorts (Extended Data 9), whereas CFP (PC2) also consistently replicated in all GNPC cohorts, although with higher heterogeneity (I^2^=86%).

Additionally, correlations between plasma and CSF PCs in paired GNPC samples (N=1,689) were negligible, indicating that protein composition is governed by different latent factors in the peripheral circulation and the CNS (Supp. Fig. 12).

### CSF turnover reference proteins improve biomarker accuracy

Given the strong influence of PC1 on brain-derived molecules, we hypothesized that identifying optimal reference markers adjusting for this trend would lead to improved AD biomarker accuracy and interpretability. We screened the Somascan dataset based on the ability of each somamer to reduce inflation in p-tau and Aβ42 signatures and improve prediction of MCI to AD-dementia conversion after residual adjustment of AD biomarker levels. The somamers with strongest PC1 correlations (based on random-effect meta-analysis derived beta coefficients) consistently reduced the inflation factor (λ) of p-tau and Aβ42 proteomic signatures (λ=∞ for p-tau; λ=62.3 for Aβ42, before adjustment), and improved 2-year MCI-to-AD conversion prediction (mean AUC=0.824 before adjustment), while those with strong PC2 correlations were largely ineffective (Fig. 5a). We report 35 somamers that consistently outperformed PC1 as adjustment variables, and were not associated to MCI to AD-dementia progression or syndromic status after Bonferroni correction (Supp. Table 8). This provides a flexible framework for researchers to select appropriate reference proteins to correct for this global proteomic trend in their datasets. By ranking these markers based on their correlation with PC1, we identified OPCML (seq.15622.13) as the top reference protein.

**Figure 5.**
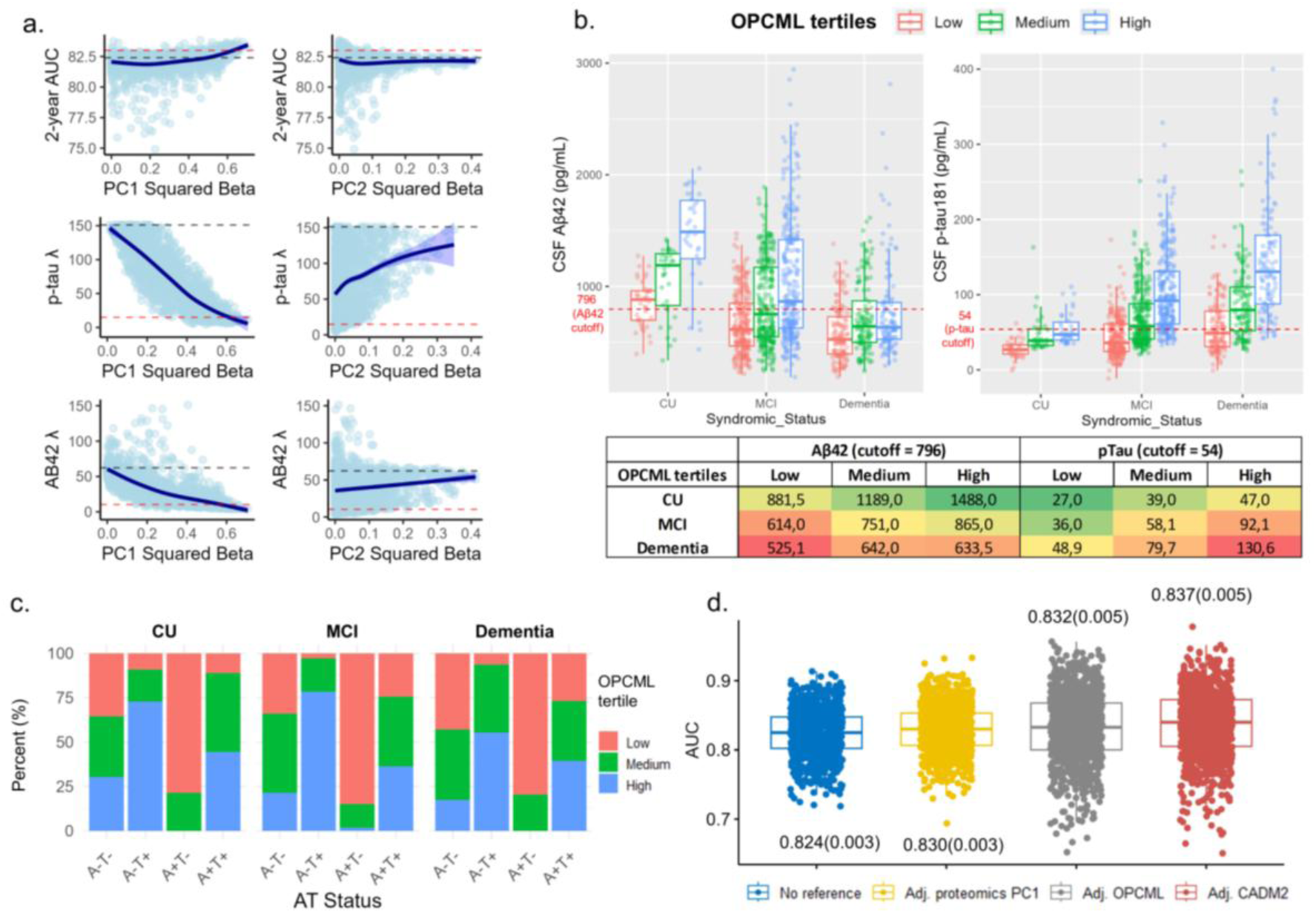
OPCML reference marker improves CSF biomarker accuracy. **a.** Mean 2-year AUC, and lambda (λ) inflation values after adjusting p-tau and Aβ42 levels by Somascan markers according to their correlation with PC1 or PC2 (measured as squared betas from lipidomics and proteomics PC counterpart random-effects meta-analysis in the ACE CSF cohort). The blue lines display generalized additive model (GAM) smooth trends. **b.** Distribution of CSF p-tau181 and Aβ42 levels stratified by disease status (CU, MCI or dementia) by OPCML tertiles within each group. Median values for each group are shown below. **c.** Proportion of individuals in each OPCML tertile, stratified by syndromic status and A/T classification. **d.** Distribution of MCI to AD conversion AUCs derived from k-means clustering (100 repetitions), using age, sex, Aβ42 and p-tau181 as predictors, depending on the reference variables used to adjust Aβ42 and p-tau181 levels.

Adjusting p-tau and Aβ42 values for OPCML dramatically reduced inflation (λ = 11.4 for p-tau and 4.8 for Aβ42) and modestly increased MCI to AD predictive accuracy to a mean AUC of 0.832 (Fig. 5d). Stratification of cognitively unimpaired (CU), MCI or dementia subjects by OPCML levels further demonstrated the strong influence of CSF turnover in biomarker concentrations: median p-tau levels in the highest tertile of CU individuals (47.0 pg/mL) were nearly identical as those of the lowest tertile in subjects with dementia (48.9 pg/mL; Fig. 5b, Table 3). Interestingly, OPCML levels were associated with age in all three strata, potentially reflecting lower CSF turnover rates associated to aging^48^, while independent from sex, MMSE, education levels and *APOE* ε4 carrier status (Table 3). AT framework classification of subjects was severely impacted by OPCML, especially those in ambiguous categories (A-T+, A+T-), with a tendency for A+T- subjects (lower concentration for both AD biomarkers) belonging in the lowest OPCML tertile, and A-T+ subjects in the highest tertile, regardless of diagnostic status (Fig. 5c, Table 3).

**Table 3.** A/T classification, demographics, MMSE and education level distribution in participants stratified by clinical status and OPCML tertiles. The p-value column shows statistical significance of Kruskal-Wallis (*DF =* 2) tests for continuous variables (p-tau, Aβ42, age, MMSE and education) or chi square tests (*DF =* 2) for categorical variables (AT classification, sex and APOE ε4 carrier status). * *p<*0.05: ** *p<*0.01; \*\*\**p>*0.001; †Chi squared test p-value including all four A/T groups (*DF =* 6).

| OPCML tertiles: | SCD |  |  |  | MCI |  |  |  | Dementia |  |  |  |
| --- | --- | --- | --- | --- | --- | --- | --- | --- | --- | --- | --- | --- |
|  | Low | Medium | High | P | Low | Medium | High | P | Low | Medium | High | P |
| Sample Size | 41 | 35 | 35 | †**4.09e-04** *** | 255 | 269 | 276 | †**1.1e-66** *** | 132 | 123 | 116 | †**5.53e-18** *** |
| A-T- | 27 | 26 | 23 | 8.43e-01 | 83 | 108 | 48 | **1.12e-05** *** | 27 | 25 | 11 | **2.68e-02** * |
| A-T+ | 1 | 2 | 8 | **2.01e-02** * | 4 | 27 | 120 | **2.82e-33** *** | 3 | 18 | 29 | **3.64e-05** *** |
| A+T- | 12 | 3 | 0 | **4.10e-04** *** | 96 | 15 | 2 | **1.22e-30** *** | 48 | 11 | 0 | **1.09e-14** *** |
| A+T+ | 1 | 4 | 4 | 3.68e-01 | 72 | 119 | 106 | **2.61e-03** ** | 54 | 69 | 76 | 1.49e-01 * |
| Aβ42<br>(median; SD) | 881.5 ;201 | 1189 ;306.8 | 1488 ;423.5 | **2.18e-09** *** | 614 ;270.6 | 751 ;369.9 | 865 ;539 | **6.41e-18** *** | 525.1 ;250 | 642 ;314.9 | 633.5 ;419.8 | **3.63e-05** *** |
| p-tau181<br>(median; SD) | 27 ;11.8 | 39 ;25.6 | 47 ;20.9 | **1.34e-11** *** | 36 ;25.7 | 58.1 ;36.6 | 92.1 ;53.6 | **2.65e-56** *** | 48.9 ;34.2 | 79.7 ;44.4 | 130.6 ;75.8 | **1.75e-27** *** |
| Age<br>(mean; SD) | 57.9 ;11.9 | 63.3 ;11.9 | 66.8 ;8.6 | **0.002** ** | 71.7 ;8.4 | 72.7 ;7.8 | 73.6 ;7.6 | **0.046** * | 73.5 ;7.8 | 75.5 ;7.8 | 75.6 ;6.7 | 0.062 |
| Females (%) | 53.7 | 57.1 | 62.9 | 0.719 | 50.2 | 53.2 | 59.1 | 0.111 | 63.6 | 64.2 | 67.2 | 0.82 |
| APOE ε4 carriers<br>(%) | 20 | 25.7 | 17.1 | 0.667 | 33.2 | 35.5 | 35.9 | 0.786 | 38.5 | 41.3 | 41.7 | 0.847 |
| MMSE<br>(mean; SD) | 29.2 ;1.7 | 29.4 ;0.7 | 29.4 ;0.9 | 0.805 | 25.6 ;3.4 | 25.8 ;3.2 | 25.9 ;2.9 | 0.799 | 20.3 ;4.5 | 21.2 ;4.6 | 20.3 ;4.5 | 0.159 |
| Education<br>(mean; SD) | 13.3 ;4.1 | 12.7 ;4 | 14.4 ;4.4 | 0.344 | 8.6 ;4.4 | 8.5 ;4.6 | 7.9 ;4.2 | 0.109 | 7 ;3.7 | 7 ;4.2 | 6.3 ;4.2 | 0.213 |

### CSF proteomic signature of AD pathology: Separating the gold from the dross

Finally, we assessed the utility of applying a reference marker for the determination of CSF p-tau and Aβ42 proteomic signatures. To maximize protein coverage, we used the complete set of Somascan 7K measurements (instead of only highly reproducible somamers, see methods). The standard model (model.1) was adjusted by age, sex, biomarker measurement technique, QAlb index and sample storage time, whereas model.2 incorporated OPCML to model.1, and model.3 incorporated OPCML and *APOE* adjustment (number of ε4 and ε2 alleles). Differentially abundant proteins (DAPs) were considered AD-increased when they showed a positive association with p-tau and a negative association to Aβ42, while AD-decreased DAPs were those associated in the opposite direction. We used the ACE CSF cohort as discovery (N=1,221), the Knight ADRC cohort^28^ as replication (N=1,073; Supp. Table 6), and meta-analysed results. As a validation strategy, we examined the overlap between our detected DAPs and those identified for ADAD mutation carriers in the DIAN cohort^12^.

In model.1, there was a negligible overlap with ADAD proteins, and results resembled the PC1 signature, with proteins such as OPCML or CADM2 among the most upregulated, and proteins such as BTK or AZU1 among the most downregulated (Fig. 6, Supp. Table 9-10). OPCML adjustment (model.2) revealed a more plausible signature, with a dominant overlap of p-tau upregulated, Aβ42 downregulated proteins (Fig. 6a). Z-scores were strongly correlated between both cohorts, with both AD biomarkers showing inverse correlations coefficients (Extended Data 10). We replicated (p<0.05) 416/458 (90.8%) OPCML-adjusted associations for Aβ42, and 719/786 (91.5%) for p-tau in the Knight ADRC cohort (Supp. Table 9). Furthermore, of the 350 DAPs detected in the ACE CSF cohort, 322 (92%) replicated in the Knight ADRC cohort (Fig. 6b, Supp. Table. 10). On the other hand, 72 somamers were Bonferroni-significant DAPs only in the Knight ADRC cohort. Interestingly, non-ADAD DAPs largely consisted of proteins identified as *APOE*-modulated^31^, with *APOE* ε4 upregulated proteins identified as AD-increased in our analysis, and vice versa (Fig. 6a-b). Consistently, these proteins lost statistical significance after *APOE* adjustment (model.3).

**Figure 6.**
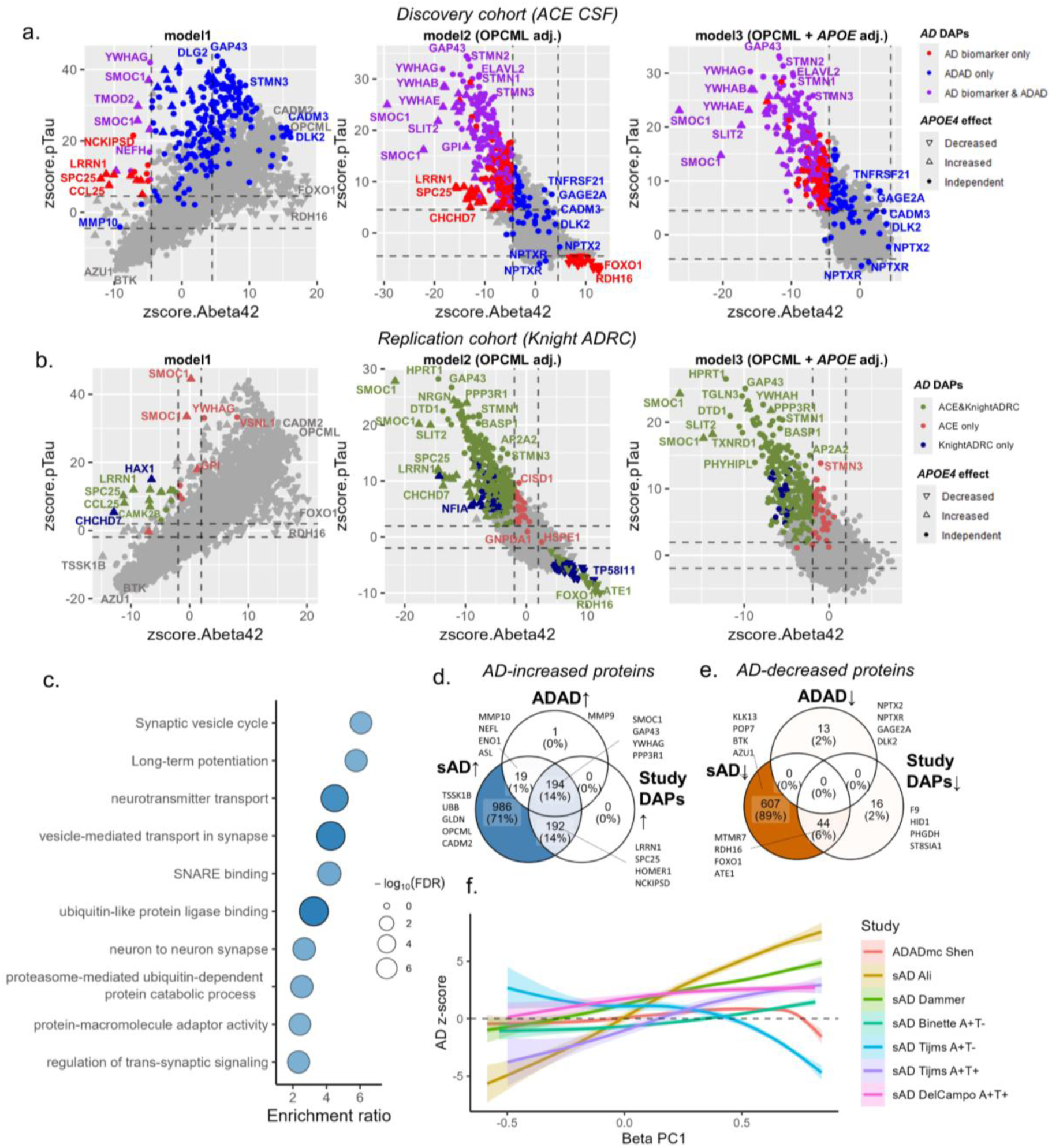
Proteomic signatures of CSF AD biomarkers. **a-b.** z-score distribution of the p-tau and Aβ42 proteomic signatures using model 1 (age, sex, QAlb index, biomarker measurement technique [ELISA/Lumipulse], sample storage time, and baseline MMSE), model 2 (model1 + OPCML) or model3 (model1 + OPCML + *APOE* ε4 and ε2 allelic dosage), in the ACE CSF and Knight ADRC cohorts, respectively. Dashed lines represent the z-score thresholds for Bonferroni and nominal significance, respectively. **c.** Top ten FDR significant GO and KEGG terms after ORA enrichment of AD-increased DAPs. **d.** Overlap between AD-increased DAPs in our study, ADAD (Shen et al.) and sAD (*Ali et al.*) **e.** Overlap between AD-decreased DAPs in our study, ADAD (Shen et al.) and sAD (*Ali et al.*). **f.** GAM smooth trend showing local averages of Alzheimer’s disease (AD) association z-scores for human proteins across different studies, as a function of the PC1 beta coefficient identified in our study.

Because the proteomic signatures were highly consistent in both cohorts, we defined the final DAPs as those with Bonferroni-significant associations in the model.2 meta-analysis, and nominal significance with consistent effect directions in the discovery and replication cohorts (Supp. Table 10). This yielded a final number of 405 (386 unique proteins) AD-increased and 60 (all unique proteins) AD-decreased somamers. Top AD-increased proteins included SMOC1, SLIT2, GAP43, 14-3-3 protein subunits (e.g. YWHAG, YWHAE), and *APOE* ε4-upregulated proteins^31^ (e.g. LRRN1, SPC25), while top AD-decreased proteins mainly consisted of known *APOE* ε4-downregulated proteins (e.g. MTMR7, RDH16, FOXO1 or TCBA)^31^. Over-representation analysis (ORA) of AD-increased proteins revealed terms such as ubiquitin-like protein ligase binding, long-term potentiation, ubiquitin-like protein ligase binding and neuron to neuron synapse, while AD-decreased proteins had no FDR-significant terms enriched (Fig. 6c, Supp. Table 11).

Although we captured the majority of the known ADAD proteomic signature^12^, panoply of proteins previously associated with sAD^11^ were not significant in our analysis (Fig. 6d-e). While our approach may lack sensitivity to detect some signals (e.g. MMP10, UBB or NEFL), the identification in previous sAD studies of OPCML and CADM2 as AD-increased, or BTK and AZU1 as AD-decreased, suggested that PC1-related confounding may be widespread in CSF proteomic studies defining disease status based on CSF biomarkers (Fig. 6d-e, Supp. Table 10). To explore this, we leveraged publicly available summary statistics from several independent CSF proteomic studies of AD^11,12,14,18,19,22^ (Supp. Table 12). Proteins showing strong correlations with PC1 tended to display inflated significance, and effect directions aligned with the PC1 trend (Fig. 6f). Interestingly, in the study by *Tijms et al.*^19^, this trend was reversed when AD status was defined solely by amyloid positivity (i.e. low Aβ42), further supporting PC1-confounding as the main driver of the observed patterns.

## Discussion

Beneath the surface of CSF biomarker readouts, our data reveals a hidden layer of pathophysiological variability—captured by PC1 (CSF turnover) and PC2 (BBB integrity)—that silently confounds current studies. In both case-control and continuous trait designs, imbalance in these latent factors drives differences that can be mistakenly attributed to the phenotype of interest, producing strong yet misleading associations and obscuring true biological signals^49^. Indeed, clear associations with known, non-CSF specific, genetic modulators of the proteome (*APOE)*^29–31^ and lipidome (*FADS1/FADS2*)^32,33^, or with disease progression (MCI to AD conversion) only emerged in our data when analysing smaller, more secondary components of the variance (Table 1, Supp. Table 4), suggesting that these subtler dimensions are the most informative to capture biological or disease-specific molecular mechanisms. Our results indicate that covariate adjustment accounting for this hidden variance—either directly via reference markers such as OPCML, or through latent variable modelling (e.g. PCA or surrogate variable analysis)—effectively enhances the robustness and interpretability of CSF biomarkers. Importantly, we provide 35 reference markers to correct for these global CSF multi-omic trend (Supp. Table 8).

Beyond confirming the presence of inter-individual variability in CSF composition^25,26^, our study expanded these observations beyond proteomics datasets, resolved them into two etiological axes (CSF turnover and BBB integrity), and revealed a complex, more structured landscape of CSF dynamics. Specifically, we identify 261 (10.9%) somamers—likely blood-derived and actively transported across the choroid plexus epithelium (e.g., leptin, prolactin)—that show inverse correlations with PC1. Elucidating the molecular mechanisms and transport domains involved in this process could inform drug development and enhance CNS delivery of therapeutic agents, including anti-amyloid antibodies.

Interestingly, most genes associated with ventricular volume (*GMNC, C16orf95, GNA12, TRIOBP, MLLT10, ARHGAP20, C11orf53)* are highly expressed in cilia-abundant tissues such as the testis and the fallopian tubes^50^, hinting a mechanistic explanation of our findings. Both choroid plexus epithelial cells and ependymal cells are enriched in motile cilia projected towards the ventricular space. The directional coordinated ciliary beating in ependymal surfaces is essential for correct CSF flow across the ventricular space and spinal canal^53^. Thus, it is biologically plausible that genetic variation in these *loci* modulates ciliary function and in turn CSF dynamics, impacting CSF composition. Moreover, *C16orf95*, *AMZ1/GNA12* and *MLLT10* are also associated with idiopathic normal pressure hydrocephalus (iNPH)^54^, suggesting that CSF drainage pathways may also contribute to the PC1 phenotype.

Beyond its genetic determinants, the PC1-associated phenotype is likely influenced by physiological and environmental exposures impacting CSF turnover. Among these factors, sleep-wake cycles play a central role in modulating both CSF production and glymphatic clearance of metabolic waste in the brain parenchyma^1,2,55–57^, both peaking during deep NREM sleep. Consistent with the PC1 trend, sleep deprivation impairs molecular clearance in the human brain^58^ and is associated with increased CSF amyloid concentration^59^, while sleep has the opposite effect, decreasing levels of amyloid and tau^60^. Investigating whether sleep quality and waking time influences the PC1 phenotype in the CSF could inform study design and sample collection protocols in biomarker research.

Moreover, we found no association of PC1 with AD pathogenesis in our study (Fig. 2d-e, Table 3, Supp. Table 4), suggesting that CSF turnover rate is unrelated to AD etiology or progression. Although this finding apparently contrasts with evidence showing impaired glymphatic clearance as contributing pathway to dementia^55,61–64^, other mechanisms specifically related to glymphatic function, such as slow-wave neuronal activity, AQP4 channel function at astrocytic endfeet, perivascular macrophage activity, and arterial pulsatility^56,65–68^, may be specifically compromised in AD, playing a distinct role in disease pathogenesis.

Our findings indicate that CSF turnover rates substantially impact AD core CSF biomarker levels. Classification of subjects within the AT framework, especially those falling into ambiguous categories—A-T+ (low AD biomarker concentration), or A+T- (high AD biomarker concentration—was strongly conditioned by OPCML levels (Table 3). Concordant A/T profiles (A+T+ or A-T-) showed a more balanced OPCML distribution, likely due to containing proportionally fewer subjects with borderline biomarker levels, and because the opposing trajectories of Aβ42 and p-tau in AD probably help counterbalance CSF turnover effects. Our results indicate that subject categorization based on CSF AD core biomarkers^4,5^ inadvertently embeds PC1-related variability in the disease classification itself. This latent variability likely contributes to the superior performance of ratio-based CSF biomarkers (e.g. p-tau/Aβ42 or Aβ42/Aβ40) compared to single analyte measures for predicting amyloid PET positivity^69^. In addition to ratio-based approaches, calibrating CSF biomarkers by these latent features will likely lead to improved accuracy in AD and other neurodegenerative disorders.

This distortion in subject categorization also pervasively confounds AD CSF proteomics studies^11,12,14,18,19,22^ (Fig. 6f, Supp. Table 12), affecting both discovery and interpretation of protein associations. This bias is evident across different platforms (SomaScan, TMT-MS, and Olink) and consistent across independent cohorts, even in studies employing surrogate variable adjustment methods^11,12^. Notably, studies applying alternative phenotyping strategies showed reduced PC1-related bias. For example, *Shen et al.*^12^ exhibited the weakest confounding in a study of autosomal dominant AD (ADAD) mutation carriers. Similarly, *Binette et al.*^14^ minimized this effect by defining amyloid positivity using the Aβ42/40 ratio, tau positivity via PET imaging, and adjusting models based on average protein levels. Importantly, PC1-related confounding can also affect studies investigating one or few biomarkers, where global proteomic trends are not so evident. For this reason, in addition to incorporating reference proteins, we recommend careful definition of the AD phenotype (preferentially using PET or ratio-based CSF biomarkers) in studies researching CSF biomarker data.

By explicitly modelling CSF dynamics, we expose a curated, more genuine signature of AD pathology (Fig. 6, Supp. Table 10). Pathway analysis of these proteins revealed terms related to loss of synaptic homeostasis and dysfunctional proteostasis, two core pathophysiological mechanisms of AD^70^. Interestingly, most DAPs exhibited upregulation, a pattern consistently observed in prior studies^11–16,18,22,24^. This upregulation likely reflects intense neurological tissue reactivity to AD pathology, either via activation of ubiquitin-mediated protein degradation pathways, or by the release of synaptic proteins caused by neuronal death.

Finally, we identified a distinct subset of the sAD CSF proteome that is strongly influenced by *APOE* genotype. These proteins (e.g. SPC25, LRRN1) show high amyloid-to-tau z-score ratios relative to other DAPs, lose significance after *APOE* adjustment, and are notably absent from ADAD proteomic signatures, where disease is driven by fully penetrant familial mutations in other loci (Fig. 6)^12,15^. The widespread impact of *APOE* ε4 on the CSF proteome, confirmed here by GWAS (Table 1) and consistent with previous reports, is neither dependent on neurodegeneration nor specific to AD^30,31,71^. Determining whether these proteins are true pathogenic mediators of *APOE*-associated AD risk, or simply reflect *APOE*-related confounding, may provide crucial insight into the still elusive mechanisms of this polymorphism.

## Methods

### Study participants: ACE CSF cohort

The ACE CSF cohort consists of 1,372 CSF samples from cognitively unimpaired (CU) subjects—5 controls and 137 with subjective cognitive decline (SCD)—individuals with mild cognitive impairment (MCI), and subjects diagnosed with dementia. Lumbar puncture (LP) was voluntarily offered to (a) individuals with mild cognitive impairment (MCI) or dementia evaluated at the memory clinic^72^, (b) participants of the Fundació ACE Healthy Brain Initiative (FACEHBI) with SCD ^73^, and (c) participants of the BIOFACE study with early-onset MCI ^74^. Briefly, syndromic diagnoses were established at the ACE Alzheimer Center Barcelona memory clinic (Barcelona, Spain) by a multidisciplinary team comprising neurologists, neuropsychologists, and social workers^72^. The baseline follow-up was defined as the closest clinical evaluation to the LP. CU classification was assigned to controls or individuals with subjective cognitive decline (SCD) who showed no objective cognitive impairment in the baseline evaluation^75^. MCI diagnoses were given to patients exhibiting one or more altered cognitive domains on the neuropsychological battery of ACE (NBACE), accounting for age and education levels, with a Clinical Dementia Rating (CDR) of 0.5 ^76,77^ at baseline. Subjects with a syndromic diagnosis of dementia and a CDR of 1 or higher were classified in the dementia group. The 2011 National Institute on Aging and Alzheimer’s Association (NIA-AA) guidelines were used to establish AD diagnoses^4^. Other dementia subtypes were categorized following the guidelines of the NINDS-AIREN criteria for vascular dementia^78^, frontotemporal dementia (FTD)^79^, and the diagnostic criteria for Lewy body dementia^80^. Information detailing past or current history of comorbidities, including cerebrovascular accident, cardiopathy, diabetes, hypertension and renal insufficiency, was recorded at the baseline visit. Further clinical characterization of these individuals has been detailed elsewhere^3^.

CSF samples were collected from the LP, following consensus recommendations, centrifuged (2,000xg for 10 minutes at 4°C), aliquoted, and stored at -80°C. For determining CSF Aβ42, total tau (t-tau), and p-tau181 protein levels, an aliquot was defrosted on the day of analysis at room temperature and vortexed. These protein levels were measured using a standard Enzyme-Linked Immunosorbent Assay (ELISA) kit (Innotest β-AMYLOID (1-42), Fujirebio Europe, Göteborg, Sweden) or the Lumipulse G600II automated platform (Fujirebio Inc.). For samples measured with Lumipulse, Aβ40 levels were also available. In-house Aβ42 and p-tau CSF cutoffs have been described elsewhere^3^. For joint analysis of the CSF biomarker levels, we transformed ELISA values to their Lumipulse equivalents based on the Passing-Bablok coefficients determined in a previous publication^3^. Additionally, all CSF samples, along with paired serum extracted during the same day, underwent routine biochemical analysis including cell counts and other relevant measures such as glucose, albumin, globulins, and total CSF protein content quantification by turbidimetry (Supp. Fig. 3).

### Study participants: Global Neurodegeneration Proteomics Consortium (GNPC)

The GNPC cohort is extensively described elsewhere^27^. This cohort constitutes the largest dataset of Somascan proteomic profiles from individuals with neurodegenerative diseases (AD, FTD, Parkinson’s disease dementia, Parkinson’s disease and amyotrophic lateral sclerosis) and cognitively unimpaired controls, collected across study sites in the USA, UK, and Europe. CSF or plasma samples were obtained from each participant at a single time point, along with demographic and clinical information. All participants provided written informed consent, and each contributing study received approval from its respective institutional ethics committee.

### Study participants: Knight ADRC cohort

Sample collection and Lumipulse CSF biomarker measurements are described elsewhere^28^. The Knight ADRC study conducts extensive longitudinal assessments, including biofluid collection, annual clinical and cognitive evaluations, neuroimaging, and brain autopsies. Participants—ranging from cognitively normal individuals to those with mild dementia—undergo comprehensive evaluations involving both clinician-led interviews and collateral informant reports. All participants were cognitively normal at baseline (CDR = 0), and incident dementia cases were identified during annual follow-up visits based on clinical assessments, following NIA-AA and NINDS diagnostic criteria. Additional study details are available at knightadrc.wustl.edu.

### Lipidomics data: lipid extraction

400 μL of cerebrospinal fluid (CSF) was mixed with 300 μL of water, 800 μl HCl(1M):CH3OH 1:8 (v/v), 900 μl CHCl3, 200 μg/ml of the antioxidant 2,6-di-tert-butyl-4-methylphenol (BHT; Sigma Aldrich), 3 μl of UltimateSPLASH™ ONE internal standard mix (#330820, Avanti Polar Lipids) and 3 μl of SphingoSPLASH™ I internal standard mix (#330734, Avanti Polar Lipids). After vortexing and centrifugation, the lower organic fraction was collected and evaporated using a Savant Speedvac spd111v (Thermo Fisher Scientific) at room temperature and the remaining lipid pellet was stored at - 20°C under argon.

### Lipidomics data: mass spectrometry

Just before mass spectrometry analysis, lipid pellets were reconstituted in 100% ethanol. Lipid species were analyzed by liquid chromatography electrospray ionization tandem mass spectrometry (LC-ESI/MS/MS) on a Nexera X2 UHPLC system (Shimadzu) coupled with hybrid triple quadrupole/linear ion trap mass spectrometer (6500+ QTRAP system; AB SCIEX). Chromatographic separation was performed on a XBridge amide column (150 mm × 4.6 mm, 3.5 μm; Waters) maintained at 35°C using mobile phase A [1 mM ammonium acetate in water-acetonitrile 5:95 (v/v)] and mobile phase B [1 mM ammonium acetate in water-acetonitrile 50:50 (v/v)] in the following gradient: (0-6 min: 0% B → 6% B; 6-10 min: 6% B → 25% B; 10-11 min: 25% B → 98% B; 11-13 min: 98% B → 100% B; 13-19 min: 100% B; 19-24 min: 0% B) at a flow rate of 0.7 mL/min which was increased to 1.5 mL/min from 13 minutes onwards. SM, CE, CER, DCER, HCER, LCER were measured in positive ion mode with a product ion of 184.1, 369.4, 264.4, 266.4, 264.4 and 264.4 respectively. TAG, DAG and MAG were measured in positive ion mode with a neutral loss for one of the fatty acyl moieties. PC, LPC, PE, LPE, PG, PI and PS were measured in negative ion mode by fatty acyl fragment ions. Lipid quantification was performed by scheduled multiple reactions monitoring (MRM), the transitions being based on the neutral losses or the typical product ions as described above. The instrument parameters were as follows: Curtain Gas = 35 psi; Collision Gas = 8 a.u. (medium); IonSpray Voltage = 5500 V and −4,500 V; Temperature = 550°C; Ion Source Gas 1 = 50 psi; Ion Source Gas 2 = 60 psi; Declustering Potential = 60 V and −80 V; Entrance Potential = 10 V and −10 V; Collision Cell Exit Potential = 15 V and −15 V.

The following fatty acyl moieties were taken into account for the lipidomic analysis: 14:0, 14:1, 16:0, 16:1, 16:2, 18:0, 18:1, 18:2, 18:3, 20:0, 20:1, 20:2, 20:3, 20:4, 20:5, 22:0, 22:1, 22:2, 22:4, 22:5 and 22:6 except for TGs which considered: 16:0, 16:1, 18:0, 18:1, 18:2, 18:3, 20:3, 20:4, 20:5, 22:2, 22:3, 22:4, 22:5, 22:6.

Peak integration was performed with the MultiQuant^TM^ software version 3.0.3. Lipid species signals were corrected for isotopic contributions (calculated with Python Molmass 2019.1.1) and were quantified based on internal standard signals and adhere to the guidelines of the Lipidomics Standards Initiative (LSI) (level 2 type quantification as defined by the LSI). Correction for batch effects was performed as described elsewhere^81^.

### Lipidomics data: quality control

The targeted lipidomics dataset consisted of 2209 lipid species measured in 1360 CSF samples. Eight samples were technical duplicates, we kept the one with the lowest missingness. We then removed 176 samples due to high missingness (>50%). We imputed the missing values of lipid species next using the impute.QRILC function of the imputeLMCD R package. Following the recommended guidelines, we removed species with over 20% missingness prior to imputation, leaving us with 820 lipid species. We then log transformed log10(x+1), mean-centered and standardized the values of the lipid species. Finally, we leveraged a previous pilot experiment with 181 overlapping samples to evaluate the reproducibility of the lipid measurements. The pilot experiment was processed using the same methodology, and we evaluated the Pearson correlation coefficients (R) between samples and lipids to assess the level of reproducibility, including the set of 522 overlapping species. The distribution of R values showed many species with null or even negative values, evidencing a fraction of the measured analytes were not reproducible among the two different experiments (Supp. Fig. 13). We decided to remove species with R<0.3 to minimize noise in our dataset and to reduce the multiple testing burden.

### Somascan data

CSF proteomic profiling for the datasets used in this study was performed using the Somascan platform (Somalogic, Boulder, CO). This technology multiplexes modified DNA aptamers, known as somamers, to quantify thousands of proteins in different human biofluids, including CSF, plasma and serum. Protein levels are expressed in relative fluorescent units (RFU) and processed using the adaptive normalization by maximum likelihood (ANML) pipeline by Somalogic. Data generation, quality control, and processing are extensively described elsewhere for the ACE CSF cohort^82^, Knight ADRC^11^, and GNPC^27^. Each individual aptamer underwent log10 transformation, mean-centering and standardization (mean=0, SD=1), and outlier removal (1.5-fold IQR). Furthermore, except in the determination of p-tau and Aβ42 proteomic signatures, we kept species with good intra-platform reproducibility, using a Spearman’s rho threshold of 0.5, resulting in a final set of 2,395 aptamers^82^.

### Genetic analyses

Germline DNA was extracted from peripheral blood and genotyped by the Spanish National Center for Genotyping (CEGEN) using the Axiom 815K Spanish Biobank Array (Thermo Fisher). We performed genotype calling using Affymetrix power tools (APT) software v1.15.0, and curated calls following the Axiom data analysis workflow. We removed samples with low genotype call rates (<97%), high heterozygosity (+3SD over mean heterozygosity), discordant genetic vs reported sex, and non-European ancestry based on the 1000G European population cluster. After sample QC, we excluded variants with high missingness (>5%), low frequency (MAF<0.01), differential missingness between cases and controls (p<10^-05^), or failing the Hardy-Weinberg equilibrium test (p<10^-06^) in the control population. We also excluded samples with genotype call rates below 97% after variant QC. Genotypes were then prepared for imputation using provided scripts (HRC-1000G-check-bim.pl) and imputed using TOPMed as reference panel using the TOPMed imputation server (Michigan, USA). We used GENESIS^83^ to infer cryptic relatedness in the resulting dataset, removing individuals with family relation within the dataset for genetic analyses. Finally, we kept 8,985,498 common variants (MAF>0.01) with good imputation quality (R2) and 1252 unrelated samples for association testing. AD PRS excluded the *APOE* region and was calculated adding the genetic doses of the risk alleles of 83 SNPs identified in the latest AD GWAS, multiplied by their effect sizes (log odds ratios)^84^. As a small fraction of our cohort took part in the former study, we calculated the PRS based on effect sizes reported at stage two. Polygenic risk score (PRS) were calculated using a custom script, as described elsewhere^85^. Briefly, the PRS is computed by adding the dosage of all reported risk alleles multiplied by their respective effect sizes (betas). Genome wide associations were obtained using PLINK v2.00a3.7LM 64-bit Intel linear models, and were adjusted by age, sex and population structure (genomic PCs 1-4).

### Multi-omic PC proteomic signatures

To test the association of somamers in the Somascan panel with PC1 and PC2, we derived unadjusted linear models for the lipidomics and proteomics PC counterparts and meta-analyzed the results. The beta coefficients for each somamer were highly correlated across the lipidomics and proteomics PCs, with larger absolute betas for proteomics PCs (Supp. Fig. 7). As PC1 and PC2 captured similar cryptic components of CSF variance across both omic datasets (Fig. 2), we focused in identifying proteins demonstrating consistent associations with both lipidomics and proteomics PCs. This approach reduced experimental noise and allowed us to define a more robust multi-omic proteomic signature for PC1 and PC2. Accordingly, we used the beta values derived from random-effect meta-analysis as a proxy metric for the correlation between the individual proteins and the cryptic phenotypes masked by PC1 and PC2 (Supp. Fig. 7).

### Statistical analysis

We used R v4.1.1. to conduct statistical analyses and data processing. All reported p-values are two-sided. As part of the FACEHBI study^73^, a small group of patients had longitudinal LPs taken at different points in time. To avoid including more than one sample obtained from the same subject, we removed one of the duplicates following these criteria: In case one of the duplicates failed the lipidomics or proteomics QC, we kept the one with both omics measurements available. Otherwise, we kept the one the with longer follow-up time. We generated the principal components (PCs) of the CSF lipidomics and proteomics datasets using the princomp function in R. To this purpose, we first calculated the covariance matrix of the lipid species in our dataset allowing missing outlier values. We calculated the mean of all mean-centered and standardized protein or lipid species, referred to as mean standardized values (MSVs), as a measure of overall signal intensity. To harmonize effect directions, we aligned the PCs with the MSV parameter, so that a higher PC value reflected higher CSF abundance. QAlb index was approximated using albumin percentages extracted from the proteinogram instead of absolute concentrations, as the latter were not available. To estimate the main PCs contributing to the MSV omic signals, we ran LASSO regressions using the glmnet R package.

The initial characterization of the omic PCs was performed by fitting linear models, including each tested PC as the dependent variable, and age, sex, CSF p-tau 181 levels, CSF Aβ42 levels, AD biomarker measurement technique (ELISA or Lumipulse), CSF turbidimetry, QAlb index, BMI, sample storage time, *APOE* ε2 and ε4 allelic dosage, and the AD PRS as independent variables. Numeric variables were mean-centered and standardized. To evaluate progression to AD, we leveraged the subjects with a MCI syndromic diagnosis at baseline and available clinical follow-ups. AD converters were defined as those converting to AD-type dementia, which required a primary diagnosis of possible or probable AD, CDR≥1, GDS≥4 and a syndromic diagnosis of dementia. Time to conversion was defined as the period between the baseline visit and conversion to AD. We censored non-converters based on their last clinical follow-up, and converters to non-AD dementia based on their date of conversion. Cox survival models were run using the survival R package^86^.

To test the association of ventricular volume increasing alleles (VVIAs) with protein and lipid species, we ran linear models adjusted by age, sex, BMI and sample storage time. To validate these results, we queried the VVIAs in independent proteomic and lipidomics QTL atlases in brain, CSF and plasma (Supp. Table 6). QTL information was obtained from Yang *et al.*^41^ (plasma proteomics and metabolomics), Wang *et al.*^33^ (CSF and brain metabolomics), Western *et al.*^30^ (CSF proteomics), and Yi *et al.*^42^ (brain proteomics). To keep both strata independent, CSF proteomics and metabolomics summary statistics were recomputed to exclude overlapping ACE CSF cohort samples. Test-statistic inflation in omics datasets was computed as the median of the χ² distribution divided by the expected median of a χ² distribution, with 1 degree of freedom. In the case of GWAS summary statistics, a MAF>0.05 filter was applied prior to genomic control λ computation.

Geneset enrichment analysis (GSEA) and over-representation analysis (ORA) were performed using WebGestalt^87^. For GSEA, we weighted the aptamers based on the signed –log10 of the p-values. For ORA, we used the list of included proteins as a reference. Gene expression profiling was performed using the GTEx tissue expression atlas (https://www.gtexportal.org).

To generate reference marker-adjusted p-tau and Aβ42 levels, we fitted linear models regressing each biomarker on the reference marker. Predicted values from these models were obtained using the predict function in R and subtracted from the original biomarker measurements, yielding residualized biomarker values. To evaluate 2-year predictive performance on MCI to AD progression we performed 10-fold cross-validation on baseline MCI individuals classified as converters (those progressing to AD within two years, using the same criteria as in the survival models) and non-converters (those with at least two years of follow-up without progression). Briefly, k-fold cross-validation splits the dataset into k equally sized folds. The model is trained on k – 1 folds and tested on the remaining one. This process is repeated k times, each time using a different fold as the test set. The results are then averaged to provide a more robust estimate of model performance. We used the createFolds function in the caret R package to generate k balanced strata, and performed AUC calculations using the roc function in the pROC R package. To account for the stochastic nature of this method, we ran each models across 100 iterations, using the mean AUC to provide a more robust estimation.

To test the association of Somascan dataset (2,395 aptamers) with CSF p-tau and Aβ42 levels, our basic model (model.1) consisted of linear regressions adjusted by age, sex, biomarker measurement technique (ELISA or Lumipulse), QAlb, sample storage time, and baseline MMSE. We then tested the effect of including OPCML levels (model.2), and OPCML plus *APOE* allelic dosages (ε4 and ε2; model.3) on the proteomic signatures. To meta-analyze the discovery (ACE CSF) and replication (Knight ADRC) cohorts, we used inverse variance-weighted fixed effect estimates, computed using the meta R package^88^. Pathway analysis of resulting up-regulated and down-regulated DAPs was performed by ORA using WebGestalt.

## Data availability

The omics and patient data supporting the findings of this study are not publicly available due to ethical and legal reasons. Access to the ACE CSF cohort data is available from the corresponding author upon reasonable request and subject to applicable ethical and data protection regulations. Knight ADRC CSF AD biomarker and Somascan data access are available upon reasonable request (https://knightadrc.wustl.edu/). The harmonized GNPC data has been made available for public request by the AD Data Initiative. Members of the global research community will be able to access the metadata and place a data use request via the AD Discovery Portal (https://discover.alzheimersdata.org/). Access is contingent upon adherence to the GNPC Data Use Agreement and the Publication Policies. The GNPC V1 harmonized data set (HDS) request link can be found on the GNPC website (https://www.neuroproteome.org/harmonized-data-set-hds).

## Code availability

The code used for data analysis has been uploaded to a GitHub repository, which will be made publicly available upon publication (https://github.com/ACE-Genomics/CSF_turnover_manuscript).

## Acknowledgements

The present work was performed as part of the doctoral thesis of Pablo García-González at the University of Barcelona (Barcelona, Spain). We would like to thank the patients and controls who participated in this project. They were processed following standard operating procedures with the appropriate approval of the Ethical and Scientific Committee. All protocols of the ACE cohort were approved by the Clinical Research Ethics Commission of the Hospital Clinic, Barcelona, Spain (reference: HCB/2014/0494) in accordance with the current Spanish regulations in the field of biomedical research and the Declaration of Helsinki. Additionally, the protocols of the HARPONE project were approved by the ethics committee of the Bellvitge University Hospital, Barcelona, Spain (reference: PR067/21, 25 February 2021).

## Author contributions

P.G-G. wrote the manuscript and conducted all statistical analyses involving the ACE CSF and GNPC cohorts. P.G-G., A.C-S. and A.R. designed the study. V.F. and A.R. supervised the work. C.Y., C.W., J.T., M.L. and M.Ali performed VVIA QTL analyses in the non-ACE cohorts. M.Ali. generated AD core biomarker proteomic signatures in the Knight ADRC cohort. J.D. and J.S. were involved in lipidomic data generation. P.G-G., I.dR and C.O. were involved in genomics data generation. Somascan data processing quality control and head-to-head comparisons were performed by R.P. Core AD biomarker measurements were performed by A.O. and L.M. Participant recruitment and clinical assessment were performed by M.Alegret. M.M., and P.S. J.M., S.E. and D.H. managed the Knight ADRC cohort. All authors revised and approved the final version of the manuscript.

## Additional information

Supplementary Information is available for this paper.

Correspondence and requests for materials should be addressed to: Agustín Ruiz MD, PhD.

GNPC V1 Supplemental Member List and Affiliations:

- Charles H Adler, Mayo Clinic Arizona, Scottsdale, Arizona, USA
- Alireza Atri, Banner Sun Health Research Institute, Sun City, Arizona, USA
- Thomas G Beach, Banner Sun Health Research Institute, Sun City, Arizona, USA
- Graham Bearden, Alzheimer’s Disease Data Initiative, Kirkland, WA
- James D. Berry, Sean M. Healey and AMG Center for ALS, Neurology
- Merce Boada, Ace Alzheimer Center Barcelona, Universitat Internacional de Catalunya, 08029 Barcelona, Spain; Biomedical Research Networking Centre in Neurodegenerative Diseases (CIBERNED), National Institute of Health Carlos III, 28029 Madrid, Spain
- Merle Bode, Hertie Institute for Clinical Brain Research, Neurodegenerative Diseases, Tübingen; German Center of Neurodegenerative Diseases, Department of Neurodegenerative Diseases, Tübingen
- Bradley Boeve, Mayo Clinic, Neurology Department, Rochester, MN
- Veronica Bot, Stanford University, The Phil and Penny Knight Initiative for Brain Resilience, Stanford, CA, USA; Stanford University, Wu Tsai Neurosciences Institute, Stanford, CA, USA; Stanford University, Graduate Program in Biomedical Engineering, Stanford, CA, USA
- Hillary Bounds, Gates Ventures, Seattle, WA
- Alfredo Cabrera-Socorro, Johnson & Johnson, NS TA, Beerse, Belgium
- Amanda Fernandez Cano, Ace Alzheimer Center Barcelona, Universitat Internacional de Catalunya, 08029 Barcelona, Spain; Biomedical Research Networking Centre in Neurodegenerative Diseases (CIBERNED), National Institute of Health Carlos III, 28029 Madrid, Spain
- Kaitlin B. Casaletto, University of California, San Francisco, Neurology Department, San Francisco, CA
- Richard J Caselli, Mayo Clinic Arizona, Scottsdale, Arizona, USA
- Yike Chen, Washington University School of Medicine, Department of Psychiatry, St. Louis, 63110, MO, USA.; NeuroGenomics and Informatics Center, Washington University School of Medicine, St. Louis, 63110, MO, USA.
- Matthew H.S. Clement, Alzheimer’s Disease Data Initiative, Kirkland, WA
- Eric B. Dammer, Emory University School of Medicine, Atlanta, GA, USA; Emory University School of Medicine, Department of Biochemistry, Atlanta, GA, USA
- Sterre de Boer, Alzheimer Center Amsterdam, Neurology, Amsterdam UMC, Amsterdam, the Netherlands; Amsterdam Neuroscience, Amsterdam, the Netherlands
- Niels De Meirleir, Johnson & Johnson, NS TA, Beerse, Belgium
- Marta del Campo Milan, Barcelonaβeta Brain Research Center (BBRC), Pasqual Maragall Foundation, Barcelona, Spain; Hospital del Mar Research Institute, Barcelona, Spain
- Daisy Ding, Stanford University, The Phil and Penny Knight Initiative for Brain Resilience, Stanford, CA, USA; Stanford University, Wu Tsai Neurosciences Institute, Stanford, CA, USA; Stanford University, Graduate Program in Biomedical Engineering, Stanford, CA, USA
- Duc Duong, Emory University School of Medicine, Atlanta, GA, USA; Emory University School of Medicine, Department of Biochemistry, Atlanta, GA, USA
- Maria Victoria Fernandez, Ace Alzheimer Center Barcelona, Universitat Internacional de Catalunya, 08029 Barcelona, Spain
- Lawrence Fourgeaud, Johnson & Johnson, NS TA, La Jolla, USA
- Raquel Puerta Fuentes, Ace Alzheimer Center Barcelona, Universitat Internacional de Catalunya, 08029 Barcelona, Spain; PhD Program in Biotecnology, Faculty of Pharmacy and Food Sciences, University of Barcelona, 08028 Barcelona, Spain
- Jordan Fuller, Gates Ventures, Seattle, WA
- Su Gao, Indiana Alzheimer’s Disease Research Center, Indianapolis, IN; Indiana University School of Medicine, Department of Biostatistics & Health Data Science, Indianapolis, IN
- John Gibbons, Rush Alzheimer’s Disease Center, Department of Neurological Sciences, Chicago, IL, USA
- Pablo Garcia Gonzalez, Ace Alzheimer Center Barcelona, Universitat Internacional de Catalunya, 08029 Barcelona, Spain; Biomedical Research Networking Centre in Neurodegenerative Diseases (CIBERNED), National Institute of Health Carlos III, 28029 Madrid, Spain
- Gyujin Heo, Washington University School of Medicine, Department of Psychiatry, St. Louis, 63110, MO, USA.; NeuroGenomics and Informatics Center, Washington University School of Medicine, St. Louis, 63110, MO, USA.
- Hilary Heuer, University of California, San Francisco, Neurology Department, San Francisco, CA
- Liping Hou, Johnson & Johnson, Spring House, USA
- Yen-Ning Huang, Indiana Alzheimer’s Disease Research Center, Indianapolis, IN; Indiana University School of Medicine, Department of Radiology & Imaging Sciences, Indianapolis, IN
- Alina Isakova, Stanford University, The Phil and Penny Knight Initiative for Brain Resilience, Stanford, CA, USA
- Clifford R. Jack, Jr, Mayo Clinic, Radiology
- Emily Kogan, Johnson & Johnson, JRD DSDH, Cambridge, USA
- Jessica B Langbaum, Banner Alzheimer’s Institute, Phoenix, Arizona, USA
- Argentina Lario-Lago, University of California, San Francisco, Neurology Department, San Francisco, CA
- Shuwei Li, Johnson & Johnson, Spring House, USA
- Shiwei Liu, Indiana Alzheimer’s Disease Research Center, Indianapolis, IN; Indiana University School of Medicine, Department of Radiology & Imaging Sciences, Indianapolis, IN
- Menghan Liu, Washington University School of Medicine, Department of Psychiatry, St. Louis, 63110, MO, USA.; NeuroGenomics and Informatics Center, Washington University School of Medicine, St. Louis, 63110, MO, USA.
- Marta Marquie, Ace Alzheimer Center Barcelona, Universitat Internacional de Catalunya, 08029 Barcelona, Spain; Biomedical Research Networking Centre in Neurodegenerative Diseases (CIBERNED), National Institute of Health Carlos III, 28029 Madrid, Spain
- Caitlin P. McHugh, Alzheimer’s Disease Data Initiative, Kirkland, WA
- Martine Meyer, Johnson & Johnson, NS TA
- Silke Miller, Johnson & Johnson, NS TA, La Jolla, USA
- Elizabeth Mlynarski, Johnson & Johnson, JRD DSDH, Spring House, USA
- Diederik Moechars, Johnson & Johnson, NS TA, Beerse, Belgium
- Patricia Moran-Losada, Stanford University, The Phil and Penny Knight Initiative for Brain Resilience, Stanford, CA, USA; Stanford University, Wu Tsai Neurosciences Institute, Stanford, CA, USA; Stanford University School of Medicine, Department of Neurology and Neurological Sciences, Stanford, CA, USA
- Paige Opsahl, Gates Ventures, Seattle, WA
- Tamina Park, Indiana Alzheimer’s Disease Research Center, Indianapolis, IN; Indiana University School of Medicine, Department of Radiology & Imaging Sciences, Indianapolis, IN
- Mukta Phatak, Alzheimer’s Disease Data Initiative, Kirkland, WA
- Joni Lindbohm, MD, PhD, University College London, UCL Brain Sciences, London, UK; University of Helsinki, Clinicum, Helsinki, Finland
- Joseph Pick, Johnson & Johnson, Spring House, USA
- Yolande AL Pijnenburg, Alzheimer Center Amsterdam, Neurology Department, Amsterdam, the Netherlands; Amsterdam Neuroscience, Amsterdam, the Netherlands
- Michael Price, Michael J. Fox Foundation, New York, NY, USA
- Shannon Risacher, Indiana Alzheimer’s Disease Research Center, Indianapolis, IN; Indiana University School of Medicine, Department of Radiology & Imaging Sciences, Indianapolis, IN
- Julio C. Rojas, University of California, San Francisco, Neurology Department, San Francisco, CA
- Howard J. Rosen, University of California, San Francisco, Neurology Department, San Francisco, CA
- Tamsin Sargood, Johnson & Johnson, Global Development, UK
- Claudia Schulte, Hertie Institute for Clinical Brain Research, Neurodegenerative Diseases, Tübingen; German Center of Neurodegenerative Diseases, Department of Neurodegenerative Diseases, Tübingen
- Weiwei Schultz, Johnson & Johnson, JRD DSDH, Titusville, USA
- Geidy E Serrano, Banner Sun Health Research Institute, Sun City, Arizona, USA
- Nicholas T. Seyfried, Emory University School of Medicine, Atlanta, GA, USA; Emory University School of Medicine, Department of Neurology, Atlanta, GA, USA; Emory University School of Medicine, Department of Biochemistry, Atlanta, GA, USA
- Todd Sherer, Michael J. Fox Foundation, New York, NY, USA
- Emily Smith, Indiana Alzheimer’s Disease Research Center, Indianapolis, IN; Indiana University School of Medicine, Department of Radiology & Imaging Sciences, Indianapolis, IN
- Adam M. Staffaroni, University of California, San Francisco, Neurology Department, San Francisco, CA
- Russell H. Swerdlow, University of Kansas Alzheimer’s Disease Research Center, Kansas City, Kansas, USA; University of Kansas, Neurology, Kansas City, Kansas, USA
- Shinya Tasaki, Rush Alzheimer’s Disease Center, Department of Neurological Sciences, Chicago, IL, USA
- Charlotte Teunissen, Neurochemistry Laboratory, Neurology Department, Amsterdam, the Netherlands; Amsterdam Neuroscience, Amsterdam, the Netherlands
- Terri G. Thompson, OnPoint Scientific, Inc, San Diego, CA, USA
- Qu Tian, NIH/NIA
- Jigyasha Timsina, Washington University School of Medicine, Department of Psychiatry, St. Louis, 63110, MO, USA.; NeuroGenomics and Informatics Center, Washington University School of Medicine, St. Louis, 63110, MO, USA.
- Abolfazl Doostparast torshizi, Johnson & Johnson, Spring House, USA
- Sergi Valero, Ace Alzheimer Center Barcelona, Universitat Internacional de Catalunya, 08029 Barcelona, Spain; Biomedical Research Networking Centre in Neurodegenerative Diseases (CIBERNED), National Institute of Health Carlos III, 28029 Madrid, Spain
- Wiesje M van der Flier, Alzheimer Center Amsterdam, Neurology Department, Amsterdam, the Netherlands; Amsterdam Neuroscience, Amsterdam, the Netherlands; Epidemiology and Data Science, Amsterdam UMC
- Julia D. Webb, University of California, San Francisco, Neurology Department, San Francisco, CA
- Bryan K Woodruff, Mayo Clinic Arizona, Scottsdale, Arizona, USA
- Ying Xu, Washington University School of Medicine, Department of Psychiatry, St. Louis, 63110, MO, USA.; NeuroGenomics and Informatics Center, Washington University School of Medicine, St. Louis, 63110, MO, USA.
- Mariet A. Younkin, Mayo Clinic, Neurology, Rochester, MN

---

**Extended Data 1.**
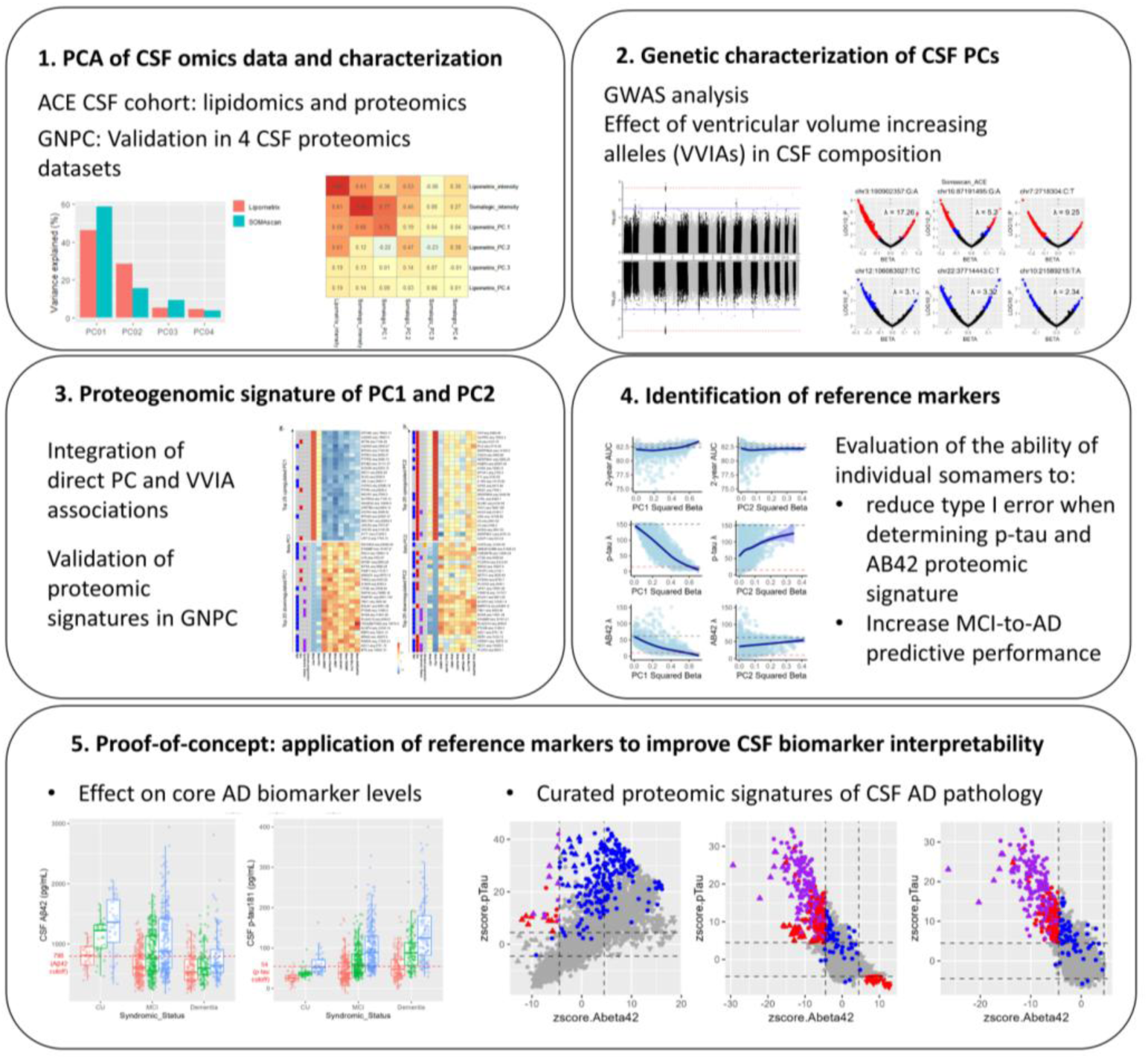
Study outline.

**Extended Data 2.**
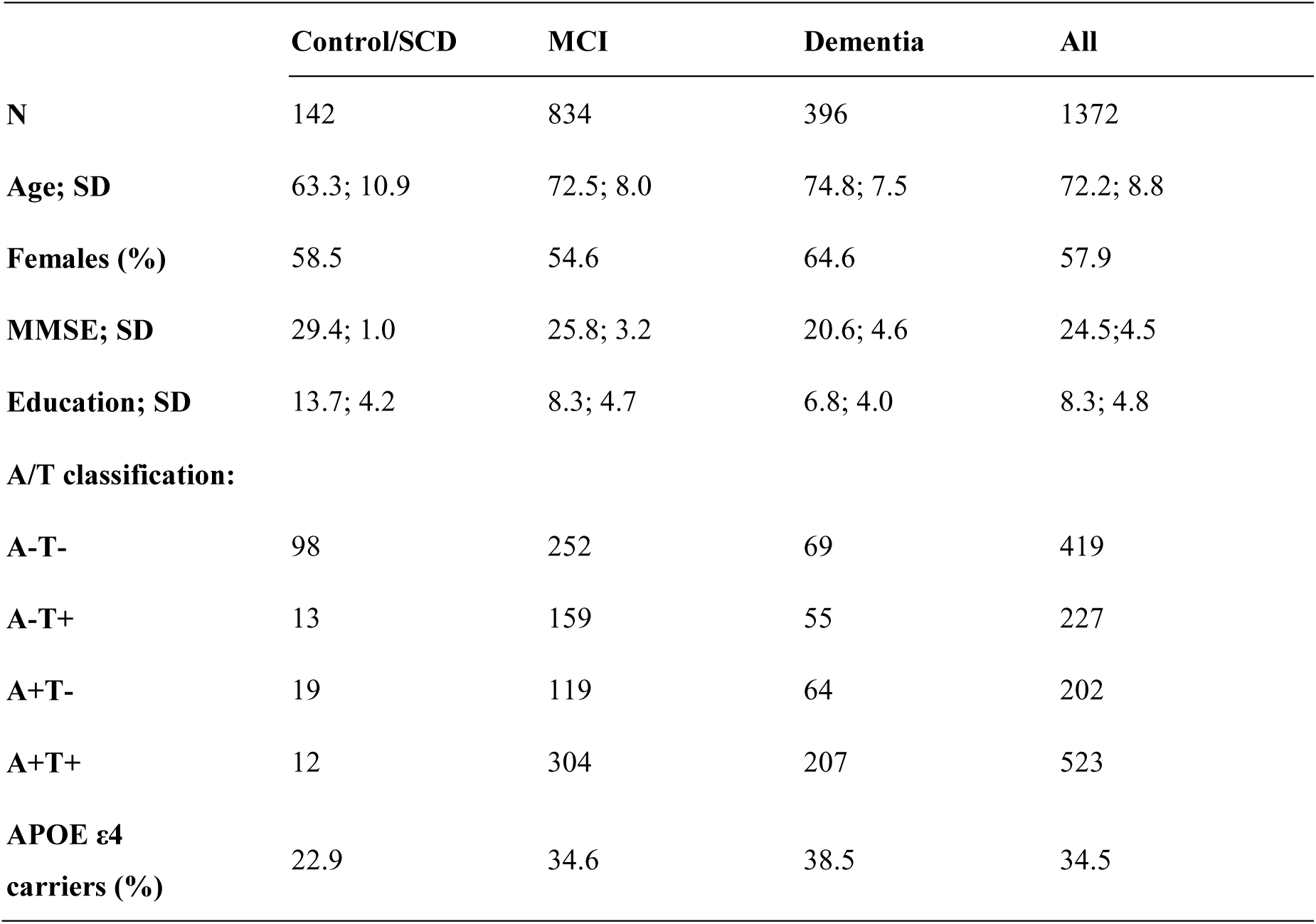
Demographics and summarized characteristics of the ACE CSF cohort. CU, cognitively unimpaired; MCI, mild cognitive impairment; Age, mean age at lumbar puncture; MMSE; mean mini mental state examination; Education, mean years of education.

**Extended Data 3.**
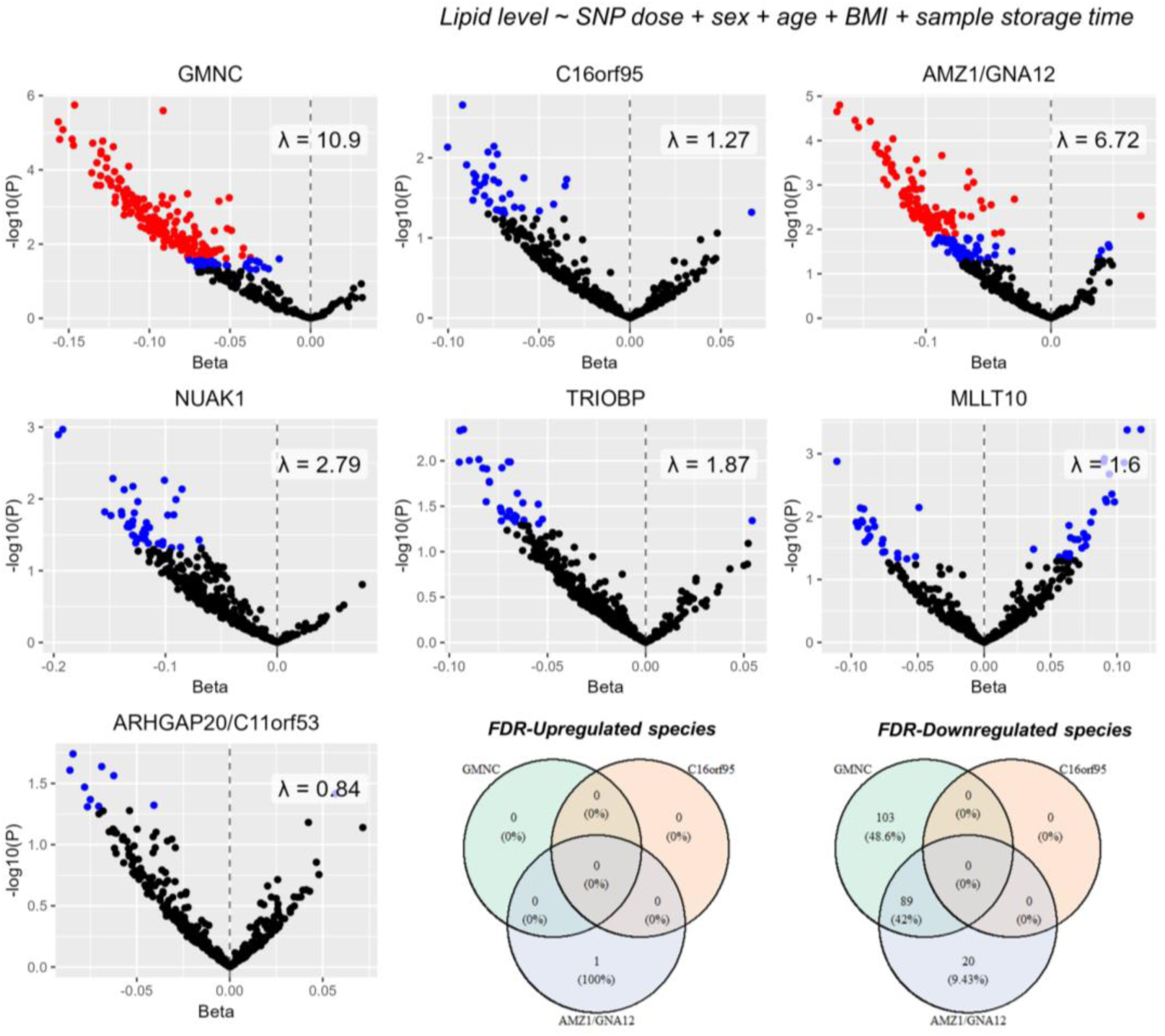
Effect of ventricular volume increasing alleles (VVIAs) on CSF lipidomics (ACE CSF cohort, *N=*1,087). Red-colored dots represent FDR significant associations (FDR_pval < 0.05), while blue dots represent nominal significant associations (*P*<0.05).

**Extended Data 4.**
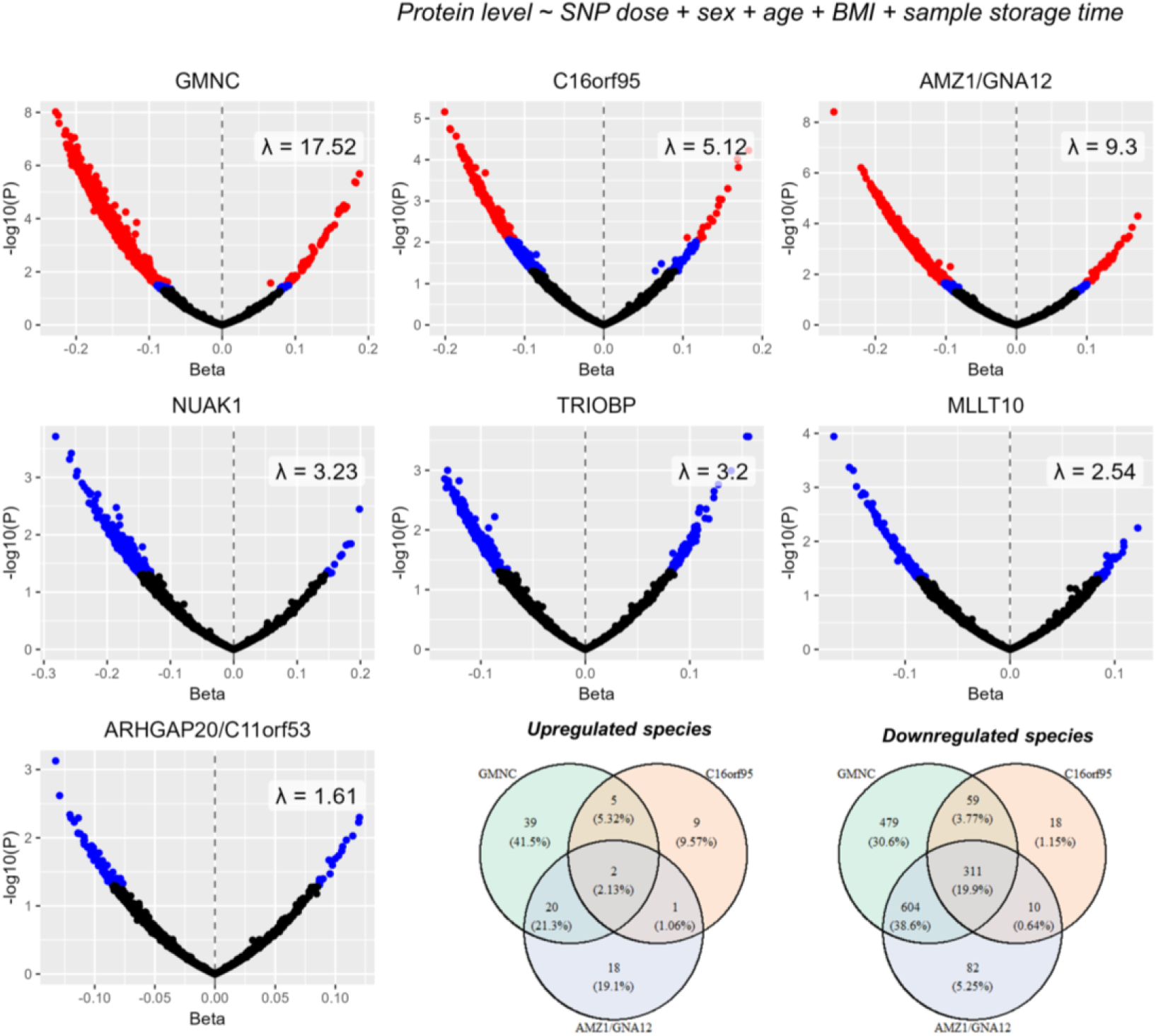
Effect of ventricular volume increasing alleles (VVIAs) on CSF proteomics (ACE CSF cohort, *N=*1,226). Red-colored dots represent FDR significant associations (FDR_pval < 0.05), while blue dots represent nominal significant associations (*P<*0.05).

**Extended Data 5.**
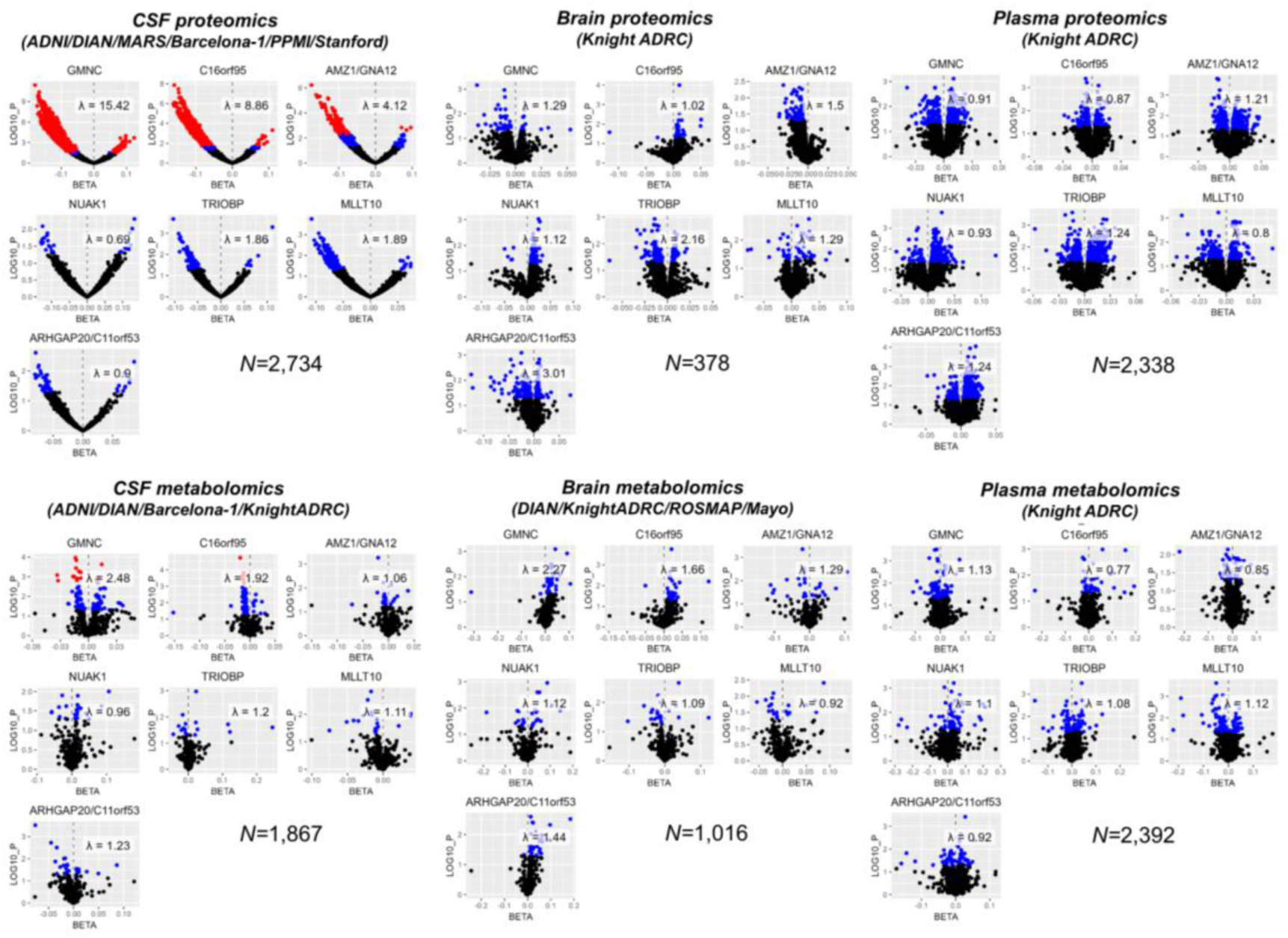
Effect of ventricular volume increasing alleles (VVIAs) in CSF plasma and brain proteomics and metabolomics data. Association results were retrieved from published QTL atlases obtained from independent cohorts (Supp. Table 6). CSF proteomics summary statistics were computed excluding samples from the ACE CSF cohort. Red-colored dots represent FDR significant associations (*FDR_pval*<0.05), while blue dots represent nominal significant associations (*P<*0.05).

**Extended Data 6.**
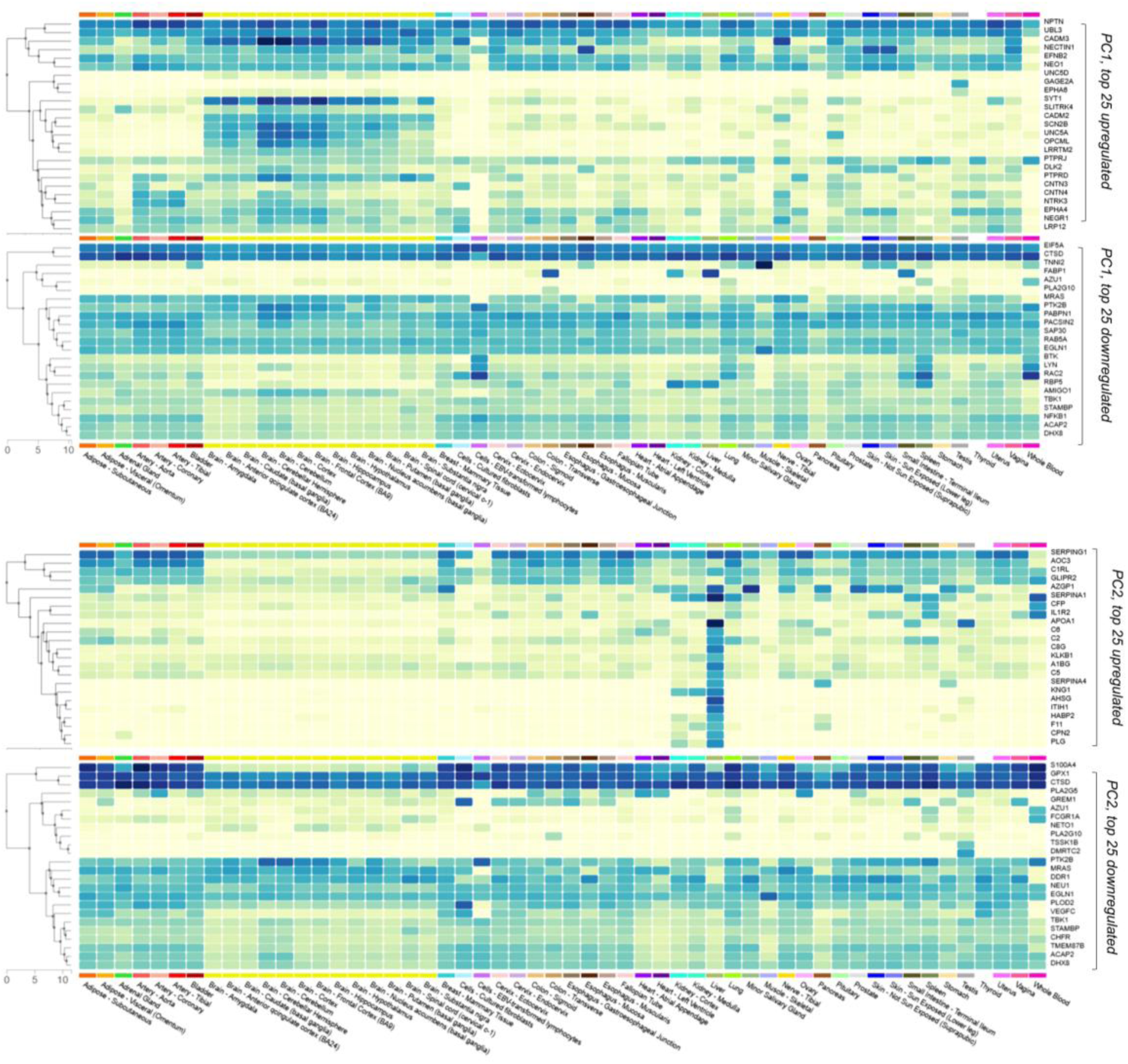
GTEx Expression Multi-Gene Query results of top 25 up- and downregulated proteins by the CSF omic PC1 and PC2 in the ACE CSF cohort. Queried 06 march 2025.

**Extended Data 7.**
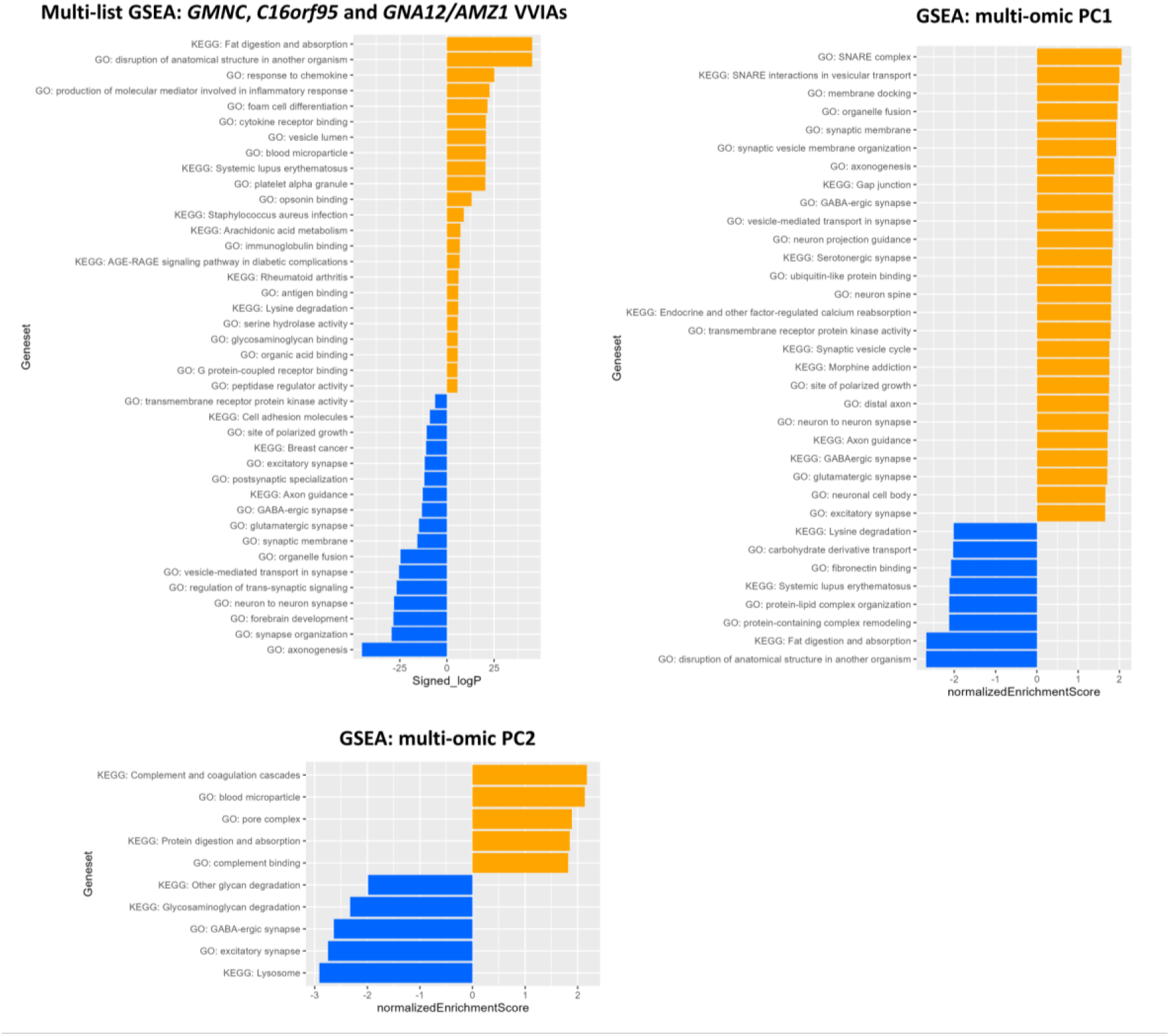
GSEA analysis for the proteomic signatures of VVIAs, PC1 and PC2 in the ACE CSF cohort. Multi-list GSEA allowed integration of VVIA signatures and was performed based on signed log p-values. In the case of PC signatures, we used the betas derived from random-effect meta-analysis of the proteomics and lipidomics datasets.

**Extended Data 8.**
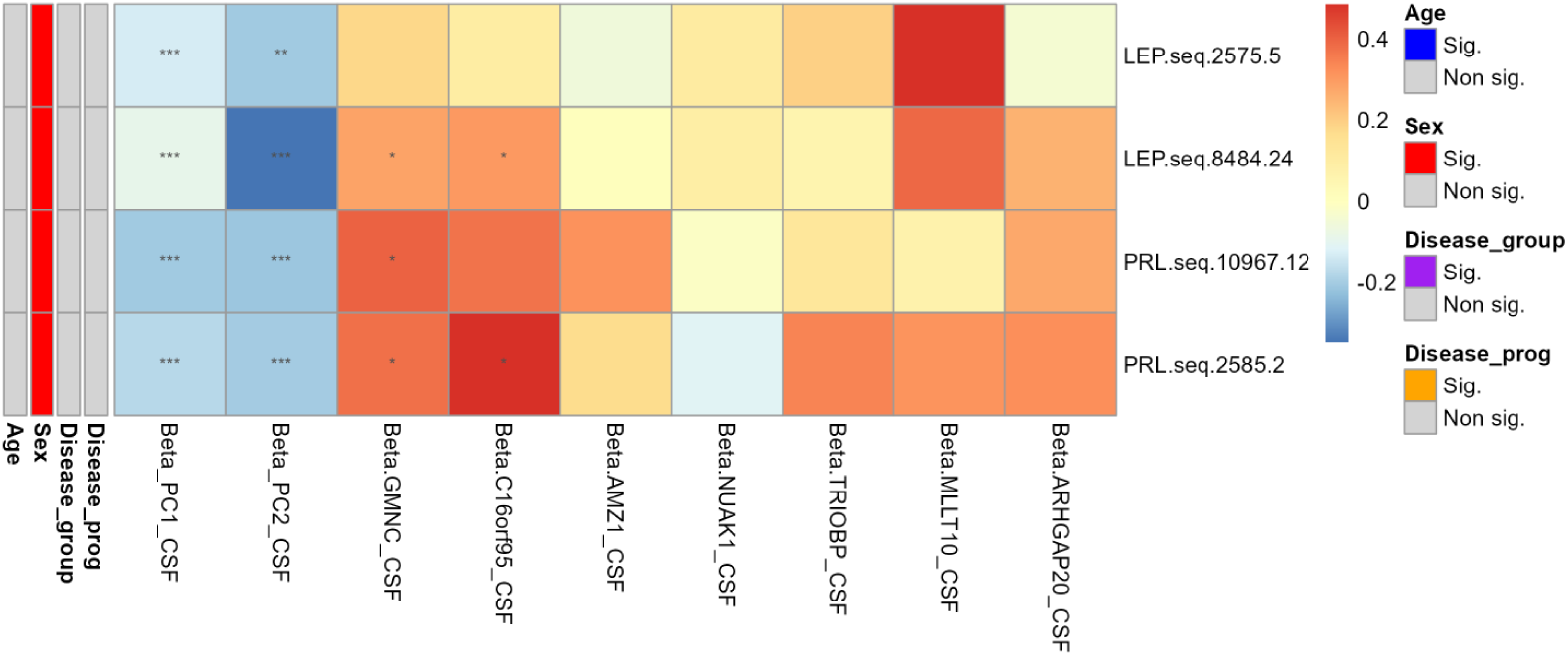
PC1, PC2 and VVIA association of the somamers binding to leptin (LEP) and prolactin (PRL) in the ACE CSF cohort (*N=*1,226).

**Extended Data 9.**
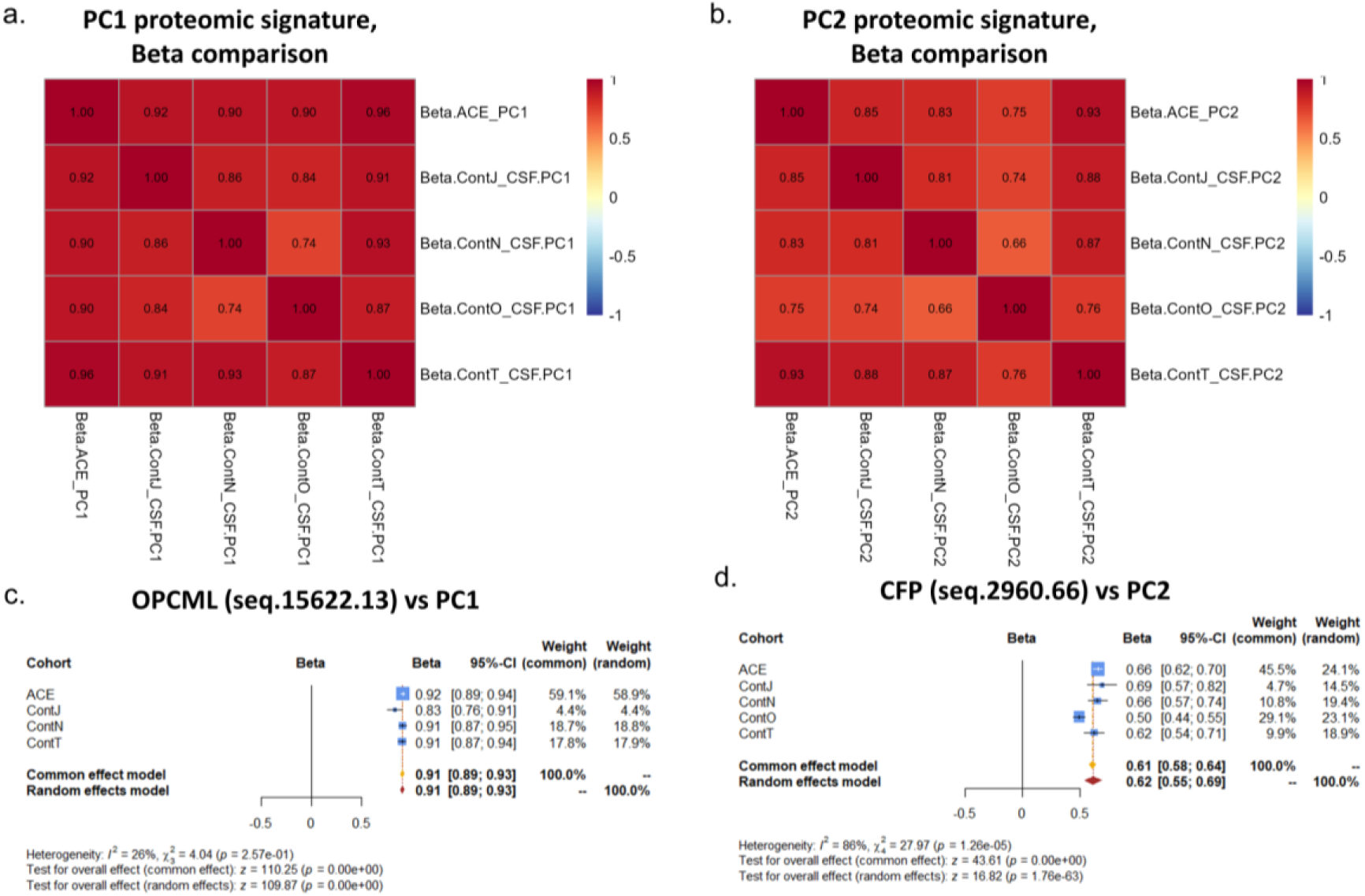
PC proteomic signature comparison across the ACE CSF cohort and GNPC cohorts. **a-b.** Pheatmap displaying Pearson correlation (R) of the Beta coefficients derived from unadjusted linear regressions fitting each individual somamer against the CSF proteomic PC1 and PC2, respectively, in the ACE CSF, and GNPC (ContJ_CSF, ContN_CSF, ContO_CSF, ContT_CSF) cohorts. **c-d.** Forest plots displaying the association of top markers for PC1 and PC2, respectively (OPCML, CFP), in the ACE CSF and GNPC cohorts.

**Extended Data 10.**
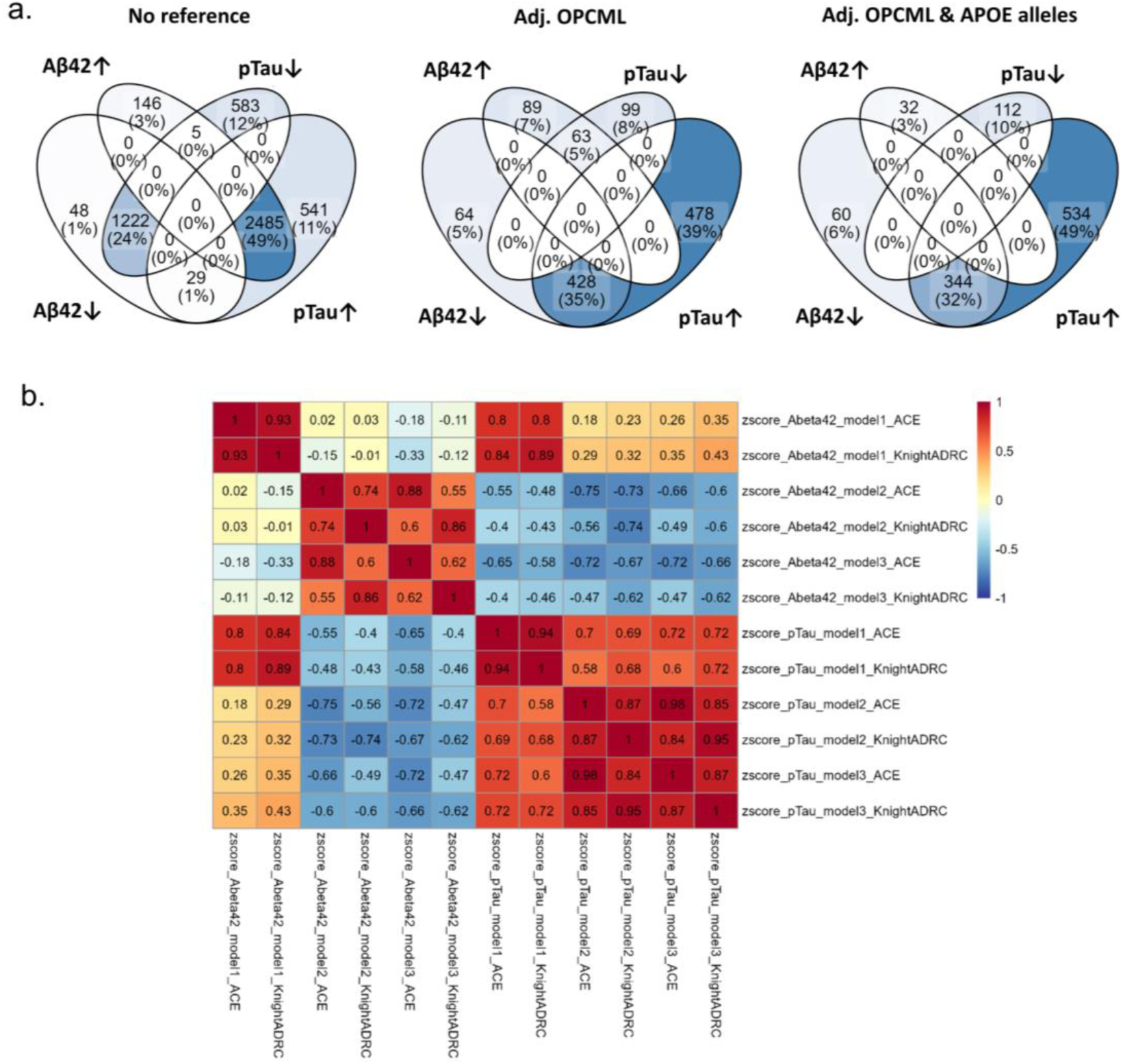
**a.** Overlap between p-tau and AB42 proteomic signatures in the different models (ACE CSF and KnightADRC meta-analysis). **b.** z-score Pearson correlation (R) between abeta42 and p-tau models in model 1 2 and 3.

