## Supplementary Figures for "CSF turnover reshapes biomarker interpretation in neurodegeneration studies"

### Slide 1
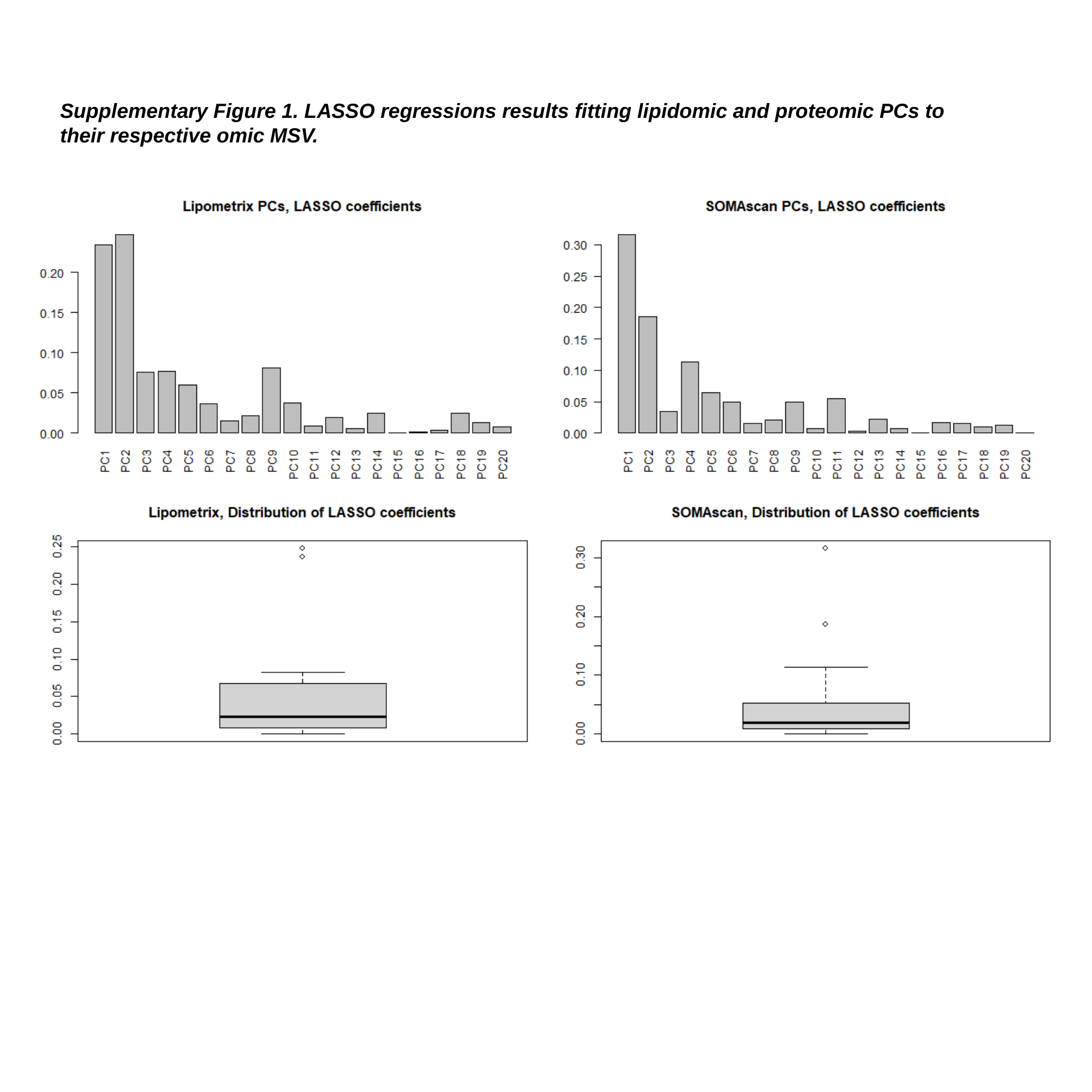

Supplementary Figure 1. LASSO regressions results fitting lipidomic and proteomic PCs to their respective omic MSV.

### Slide 2
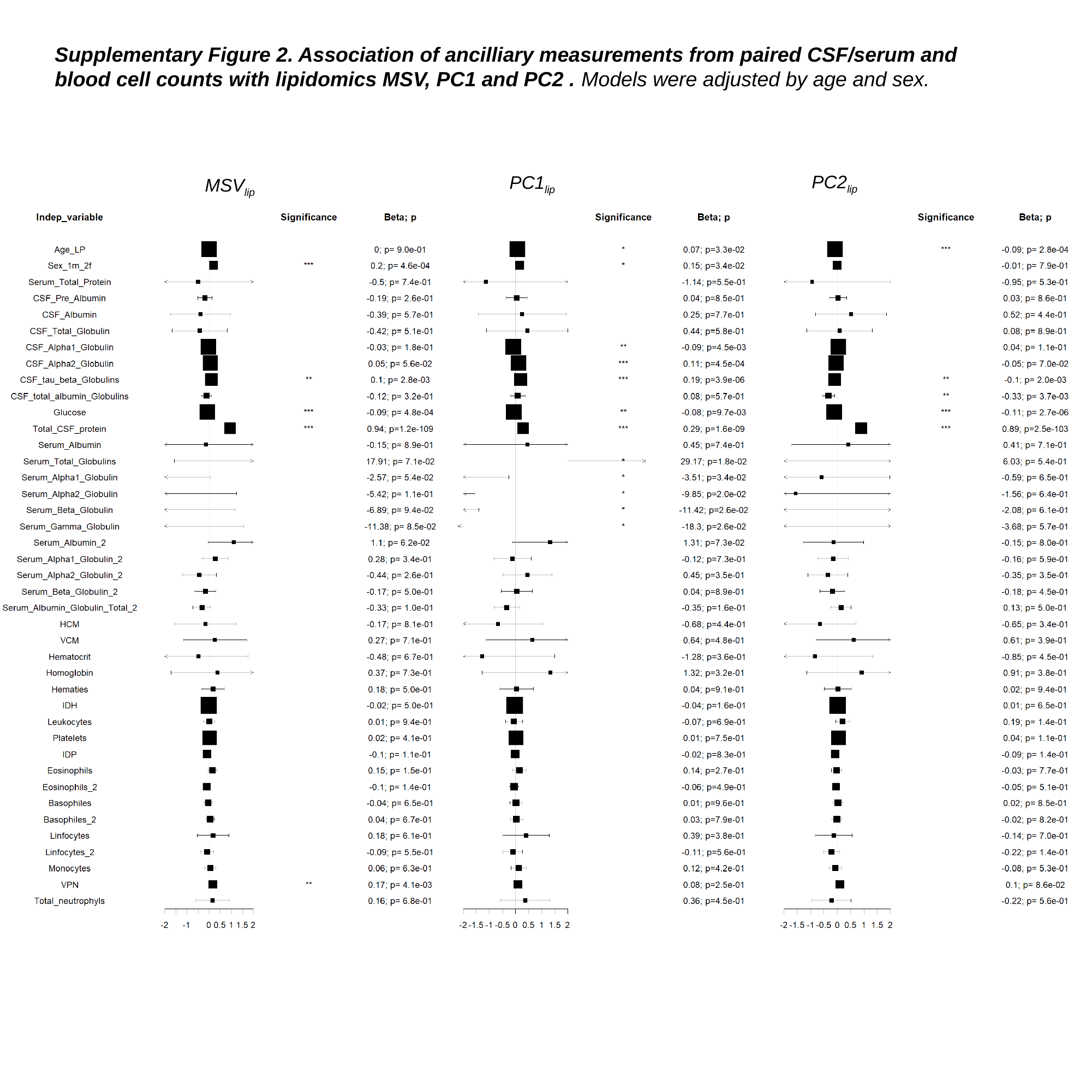

Supplementary Figure 2. Association of ancilliary measurements from paired CSF/serum and blood cell counts with lipidomics MSV, PC1 and PC2 . Models were adjusted by age and sex.
PC2_lip_
PC1_lip_
MSV_lip_

### Slide 3
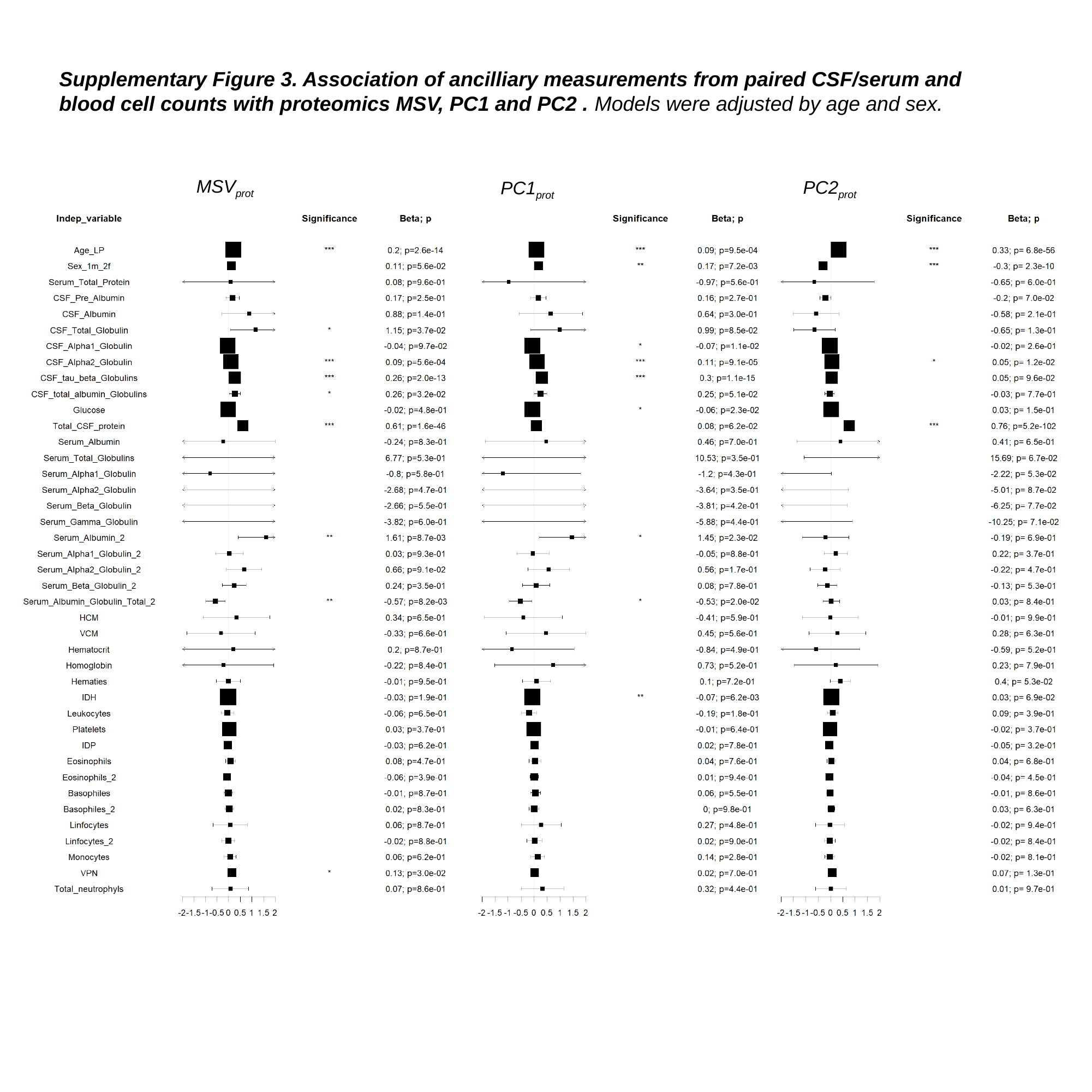

Supplementary Figure 3. Association of ancilliary measurements from paired CSF/serum and blood cell counts with proteomics MSV, PC1 and PC2 . Models were adjusted by age and sex.
MSV_prot_
PC2_prot_
PC1_prot_

### Slide 4
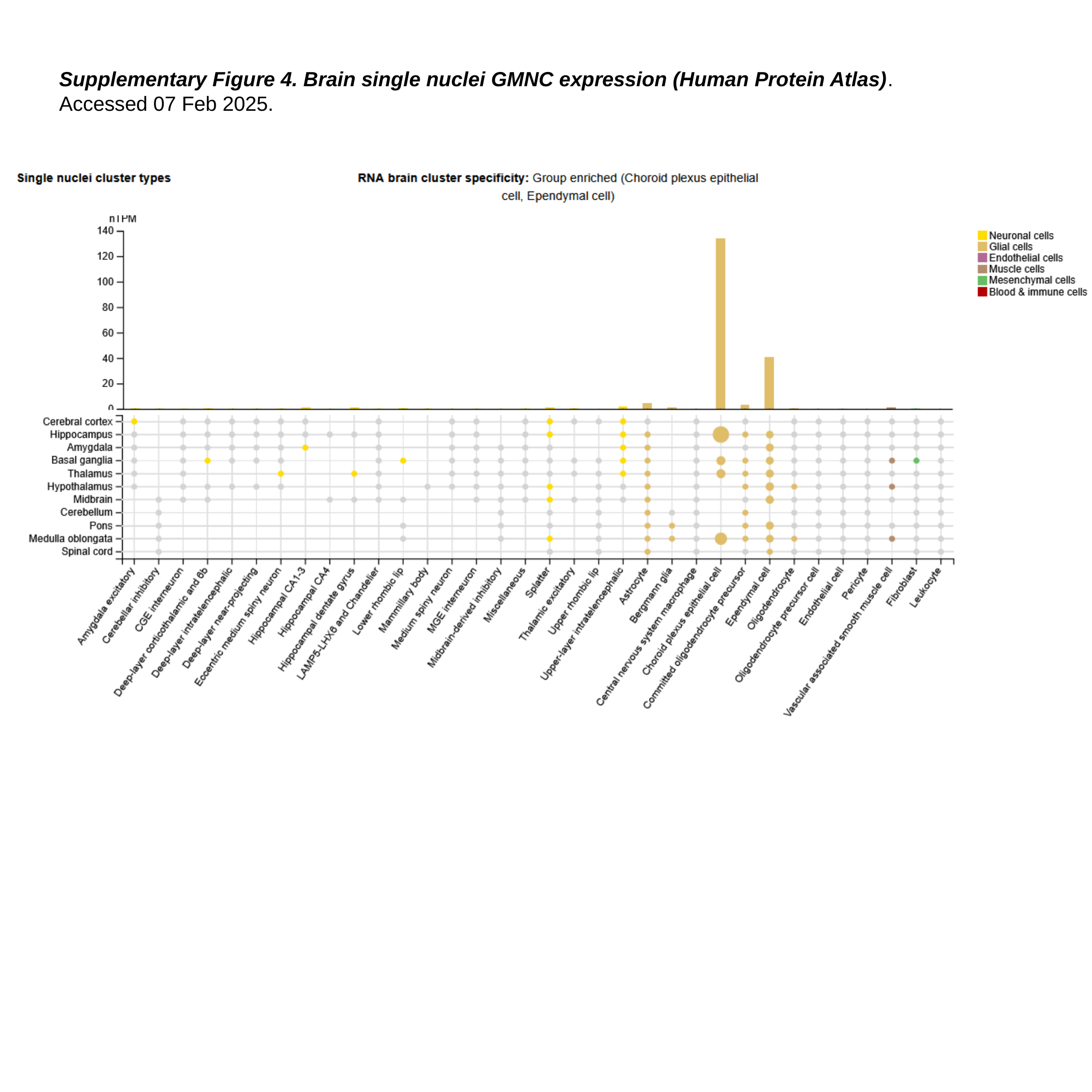

Supplementary Figure 4. Brain single nuclei GMNC expression (Human Protein Atlas). Accessed 07 Feb 2025.

### Slide 5
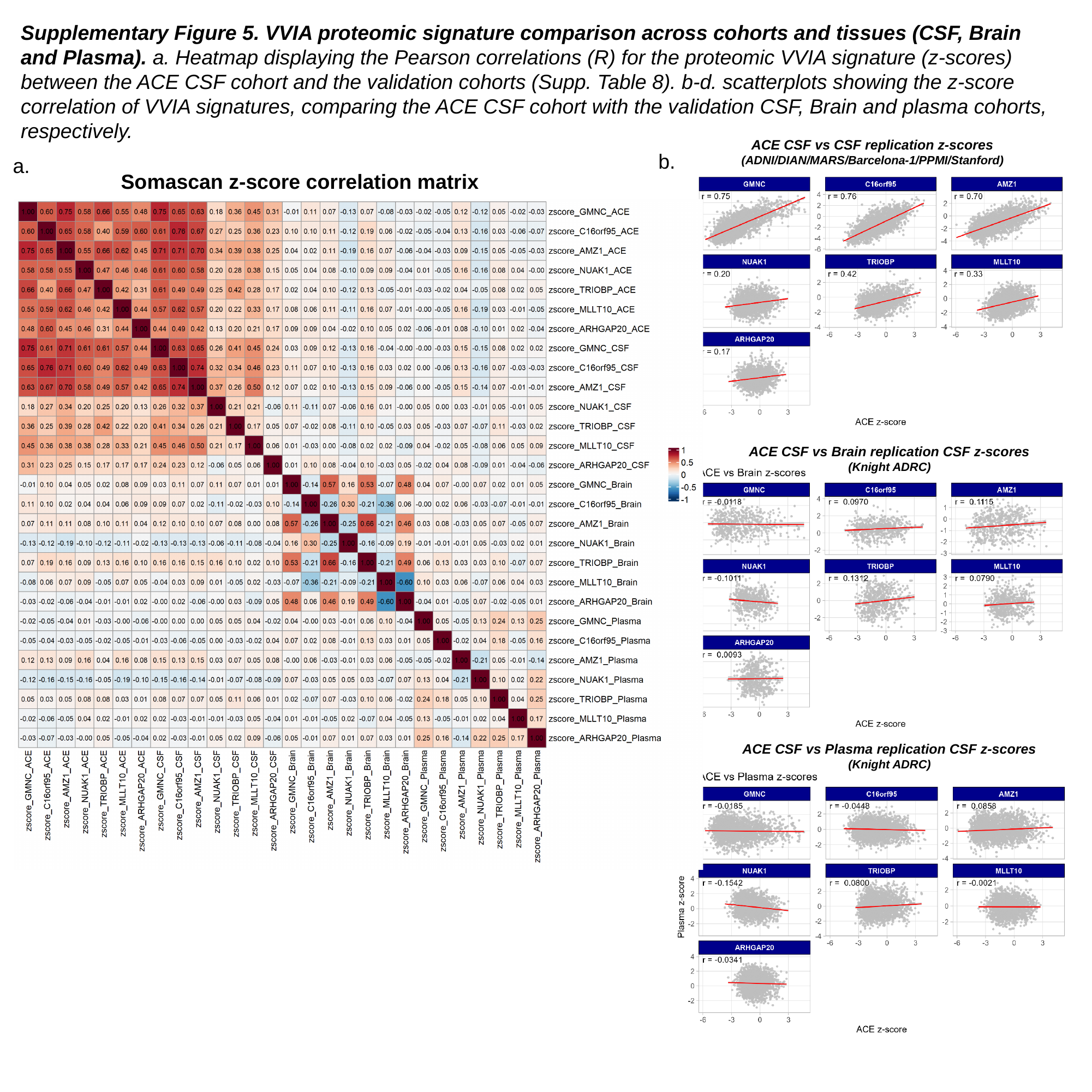

Supplementary Figure 5. VVIA proteomic signature comparison across cohorts and tissues (CSF, Brain and Plasma). a. Heatmap displaying the Pearson correlations (R) for the proteomic VVIA signature (z-scores) between the ACE CSF cohort and the validation cohorts (Supp. Table 8). b-d. scatterplots showing the z-score correlation of VVIA signatures, comparing the ACE CSF cohort with the validation CSF, Brain and plasma cohorts, respectively.
ACE CSF vs CSF replication z-scores
(ADNI/DIAN/MARS/Barcelona-1/PPMI/Stanford)
b.
a.
Somascan z-score correlation matrix
ACE CSF vs Brain replication CSF z-scores
(Knight ADRC)
c.
ACE CSF vs Plasma replication CSF z-scores
(Knight ADRC)
d.

### Slide 6
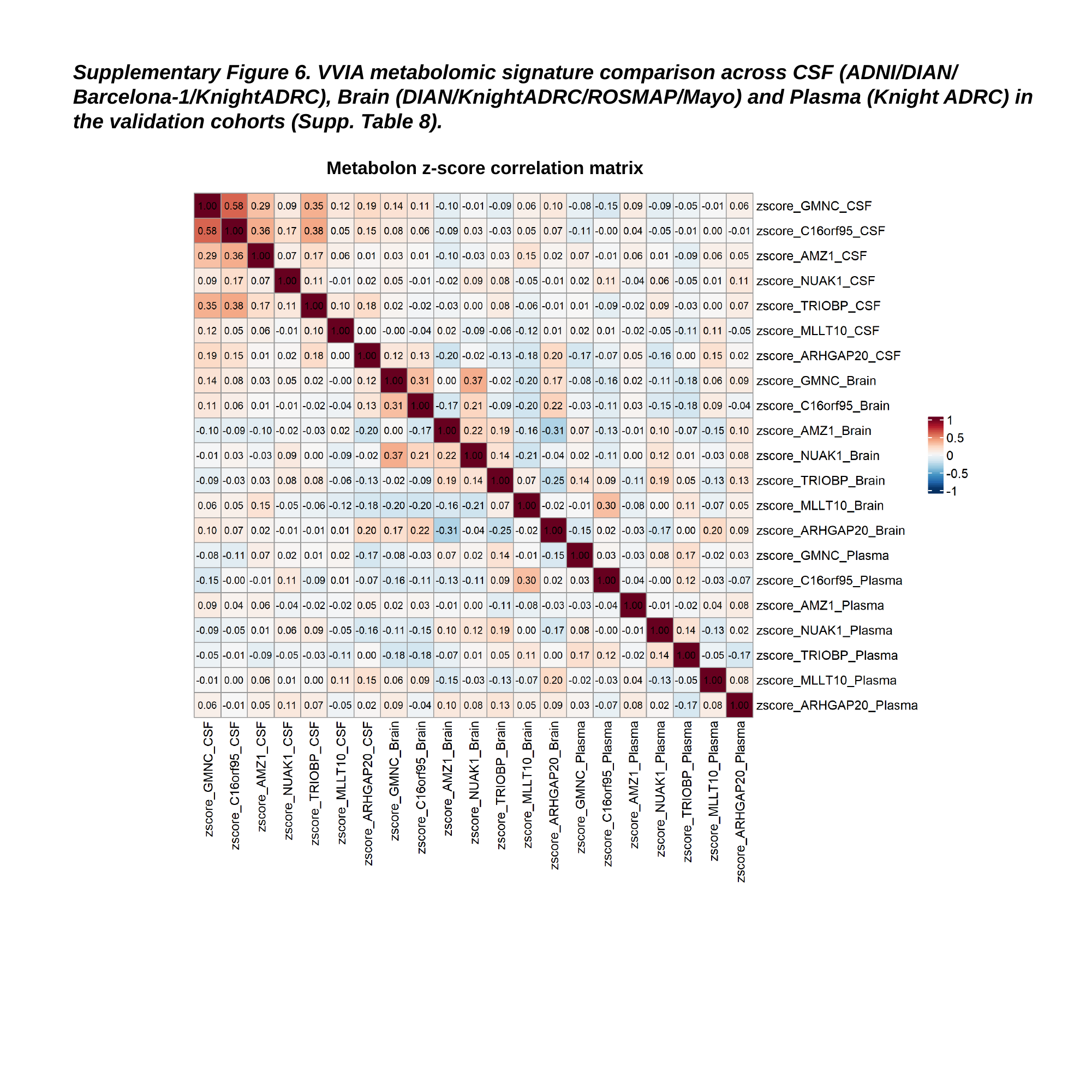

Supplementary Figure 6. VVIA metabolomic signature comparison across CSF (ADNI/DIAN/ Barcelona-1/KnightADRC), Brain (DIAN/KnightADRC/ROSMAP/Mayo) and Plasma (Knight ADRC) in the validation cohorts (Supp. Table 8).
Metabolon z-score correlation matrix

### Slide 7
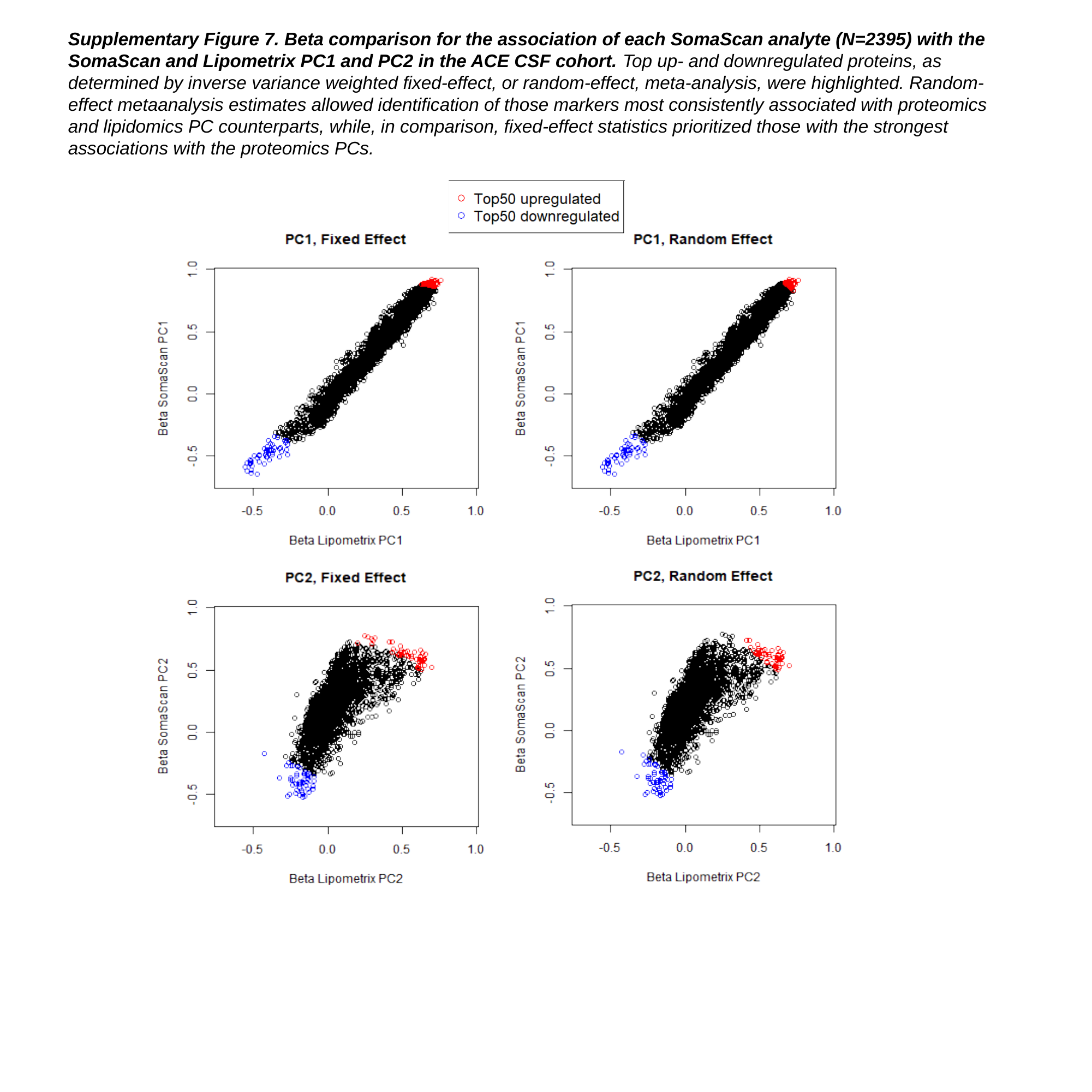

Supplementary Figure 7. Beta comparison for the association of each SomaScan analyte (N=2395) with the SomaScan and Lipometrix PC1 and PC2 in the ACE CSF cohort. Top up- and downregulated proteins, as determined by inverse variance weighted fixed-effect, or random-effect, meta-analysis, were highlighted. Random-effect metaanalysis estimates allowed identification of those markers most consistently associated with proteomics and lipidomics PC counterparts, while, in comparison, fixed-effect statistics prioritized those with the strongest associations with the proteomics PCs.

### Slide 8
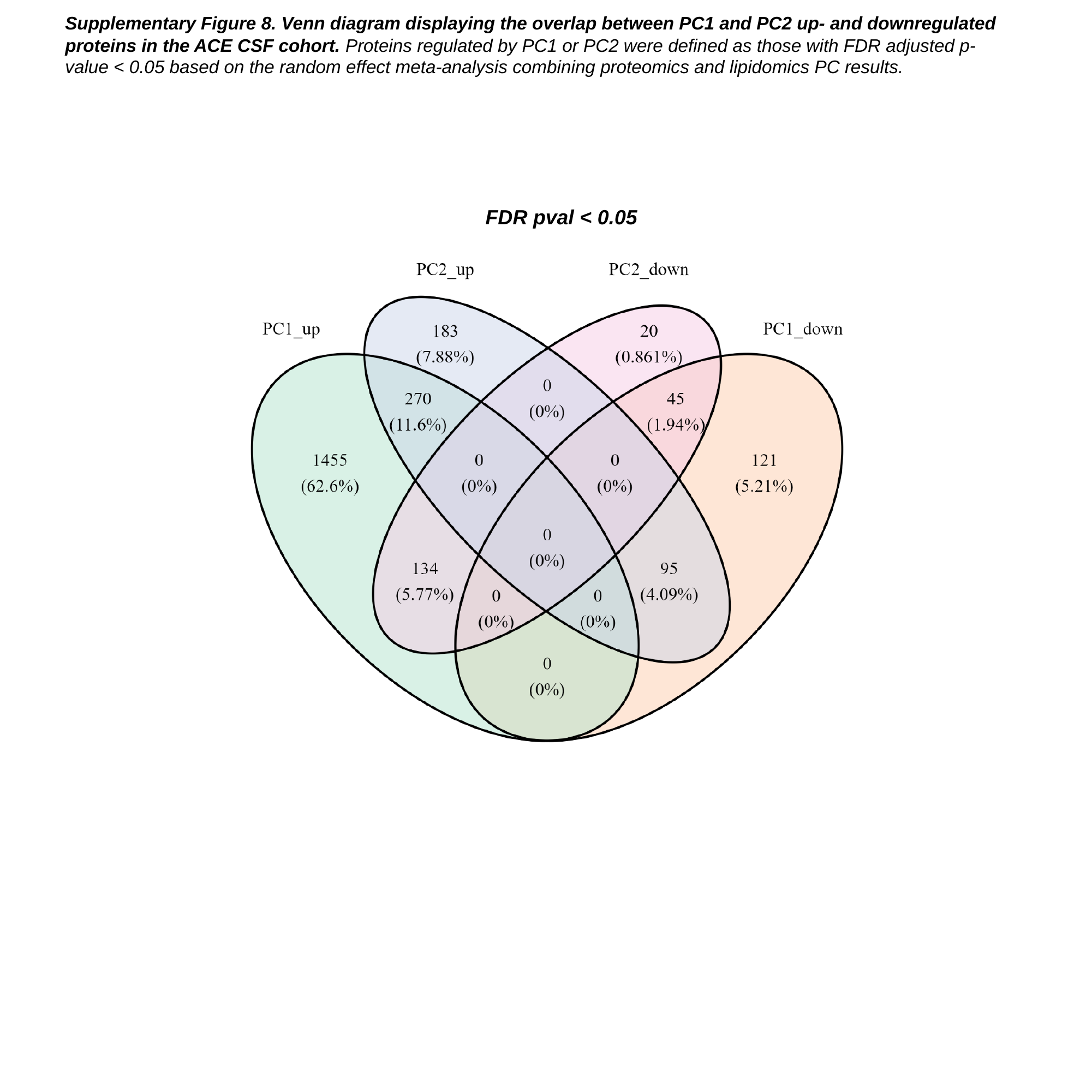

Supplementary Figure 8. Venn diagram displaying the overlap between PC1 and PC2 up- and downregulated proteins in the ACE CSF cohort. Proteins regulated by PC1 or PC2 were defined as those with FDR adjusted p-value < 0.05 based on the random effect meta-analysis combining proteomics and lipidomics PC results.
FDR pval < 0.05

### Slide 9
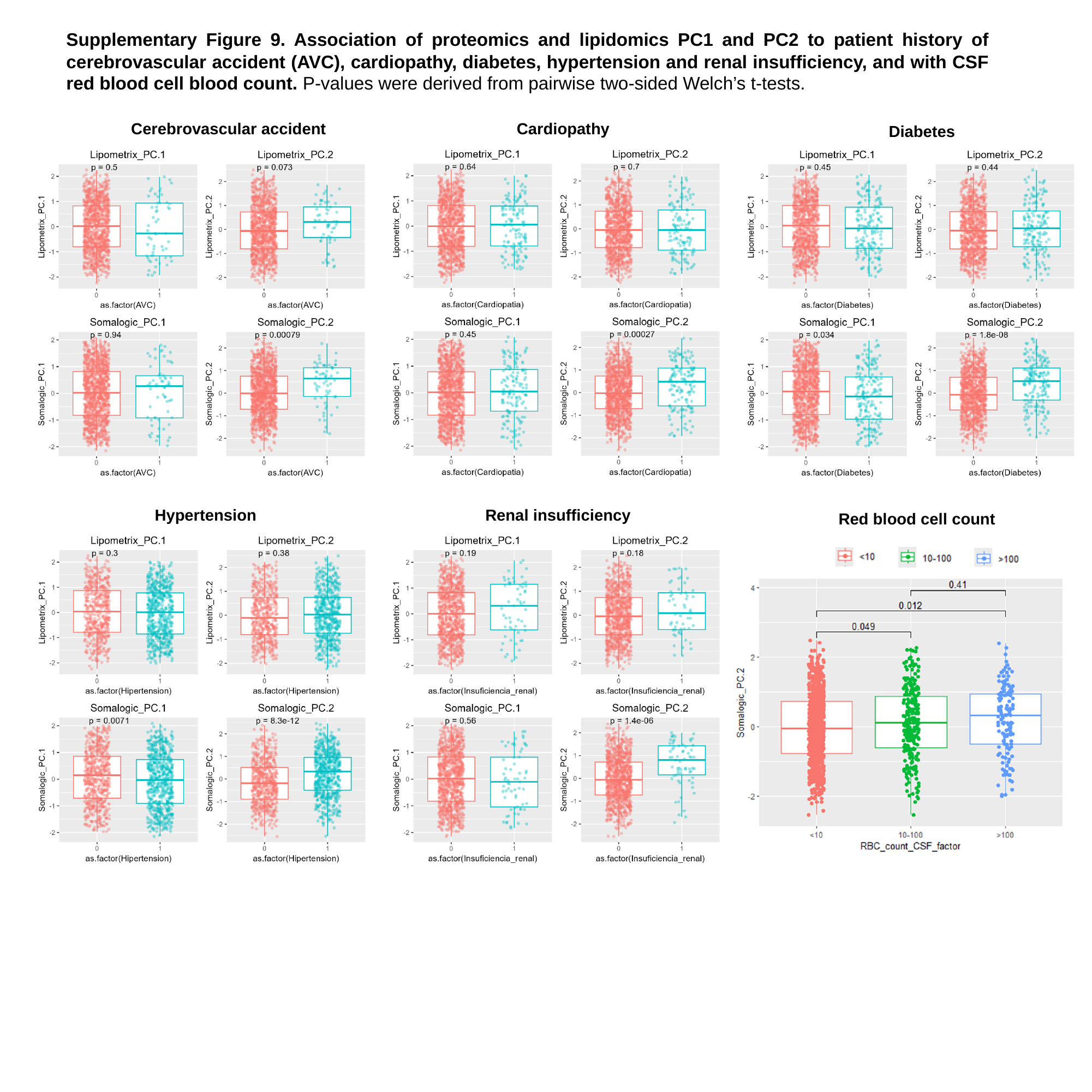

Supplementary Figure 9. Association of proteomics and lipidomics PC1 and PC2 to patient history of cerebrovascular accident (AVC), cardiopathy, diabetes, hypertension and renal insufficiency, and with CSF red blood cell blood count. P-values were derived from pairwise two-sided Welch’s t-tests.
Cerebrovascular accident
Cardiopathy
Diabetes
Hypertension
Renal insufficiency
Red blood cell count

### Slide 10
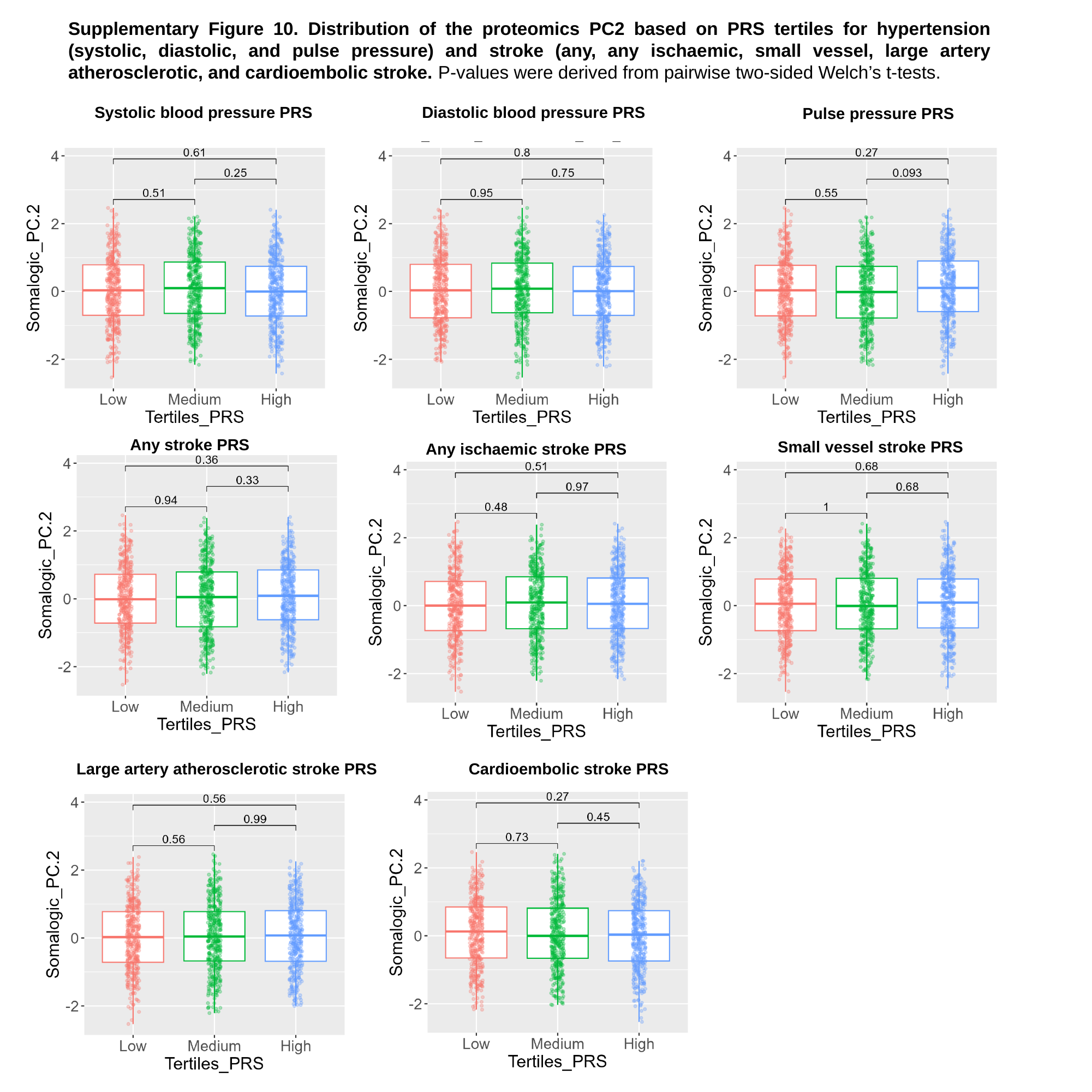

Supplementary Figure 10. Distribution of the proteomics PC2 based on PRS tertiles for hypertension (systolic, diastolic, and pulse pressure) and stroke (any, any ischaemic, small vessel, large artery atherosclerotic, and cardioembolic stroke. P-values were derived from pairwise two-sided Welch’s t-tests.
Diastolic blood pressure PRS
Systolic blood pressure PRS
Pulse pressure PRS
Any stroke PRS
Small vessel stroke PRS
Any ischaemic stroke PRS
Large artery atherosclerotic stroke PRS
Cardioembolic stroke PRS

### Slide 11
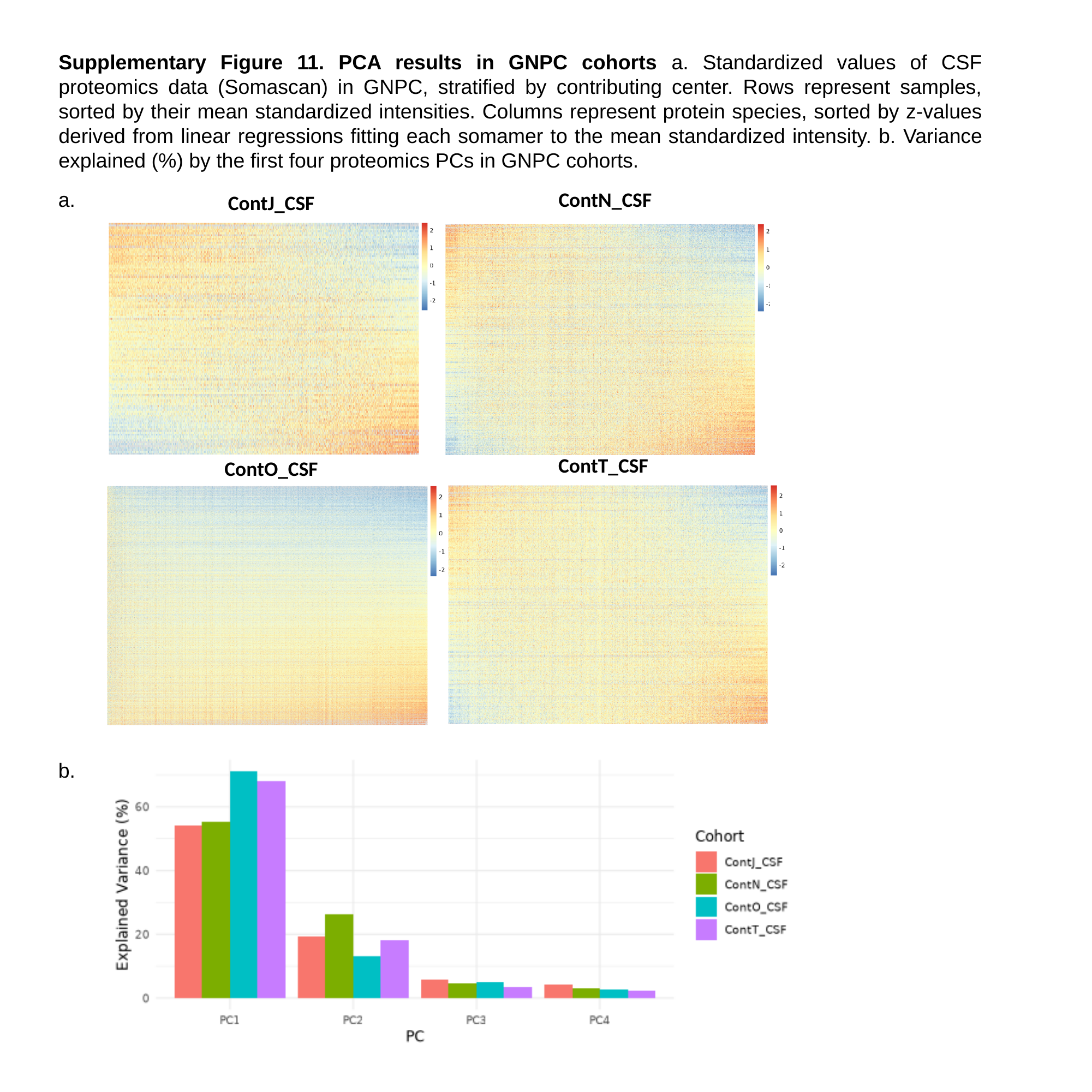

Supplementary Figure 11. PCA results in GNPC cohorts a. Standardized values of CSF proteomics data (Somascan) in GNPC, stratified by contributing center. Rows represent samples, sorted by their mean standardized intensities. Columns represent protein species, sorted by z-values derived from linear regressions fitting each somamer to the mean standardized intensity. b. Variance explained (%) by the first four proteomics PCs in GNPC cohorts.
a.
ContN_CSF
ContJ_CSF
ContT_CSF
ContO_CSF
b.

### Slide 12
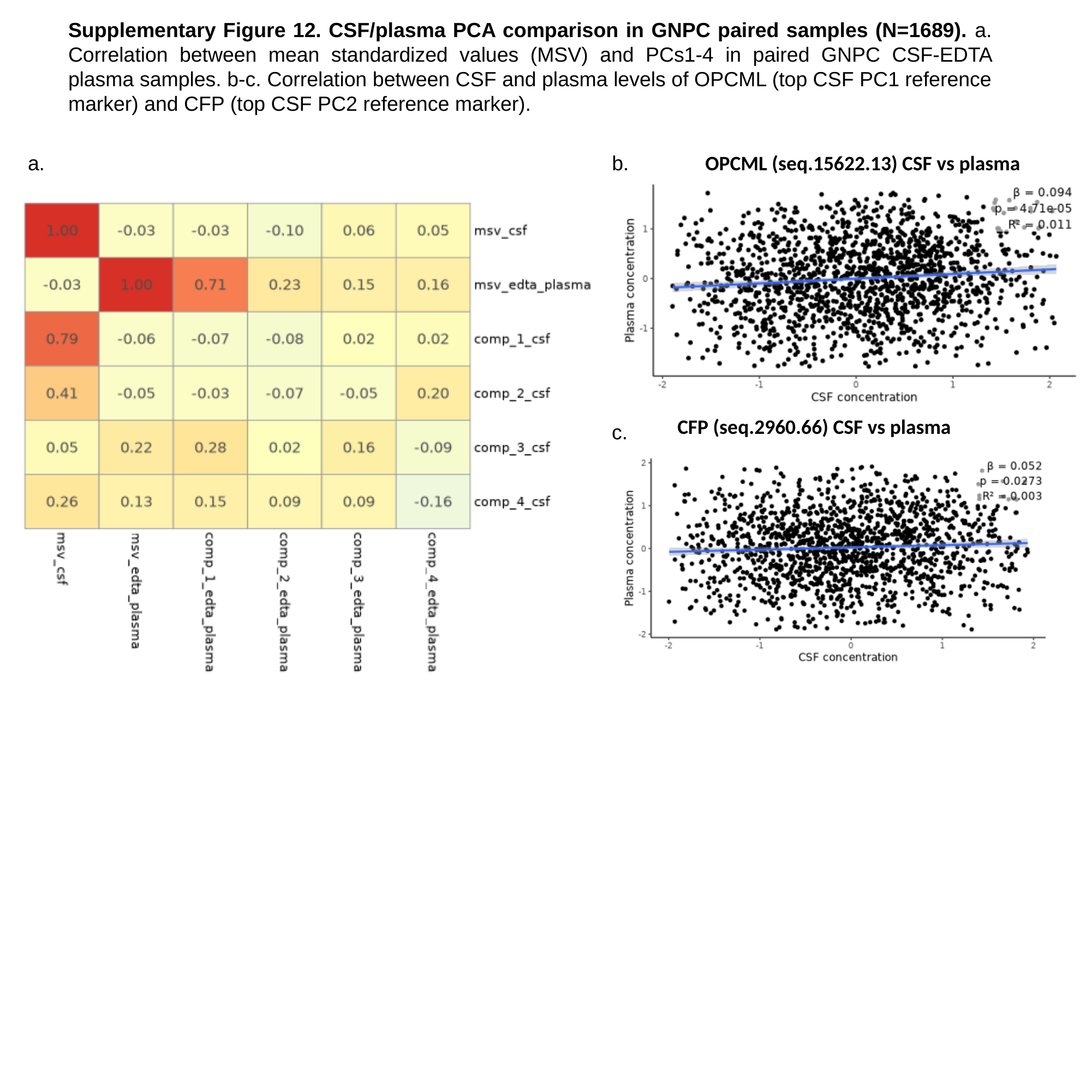

Supplementary Figure 12. CSF/plasma PCA comparison in GNPC paired samples (N=1689). a. Correlation between mean standardized values (MSV) and PCs1-4 in paired GNPC CSF-EDTA plasma samples. b-c. Correlation between CSF and plasma levels of OPCML (top CSF PC1 reference marker) and CFP (top CSF PC2 reference marker).
a.
b.
OPCML (seq.15622.13) CSF vs plasma
CFP (seq.2960.66) CSF vs plasma
c.

### Slide 13
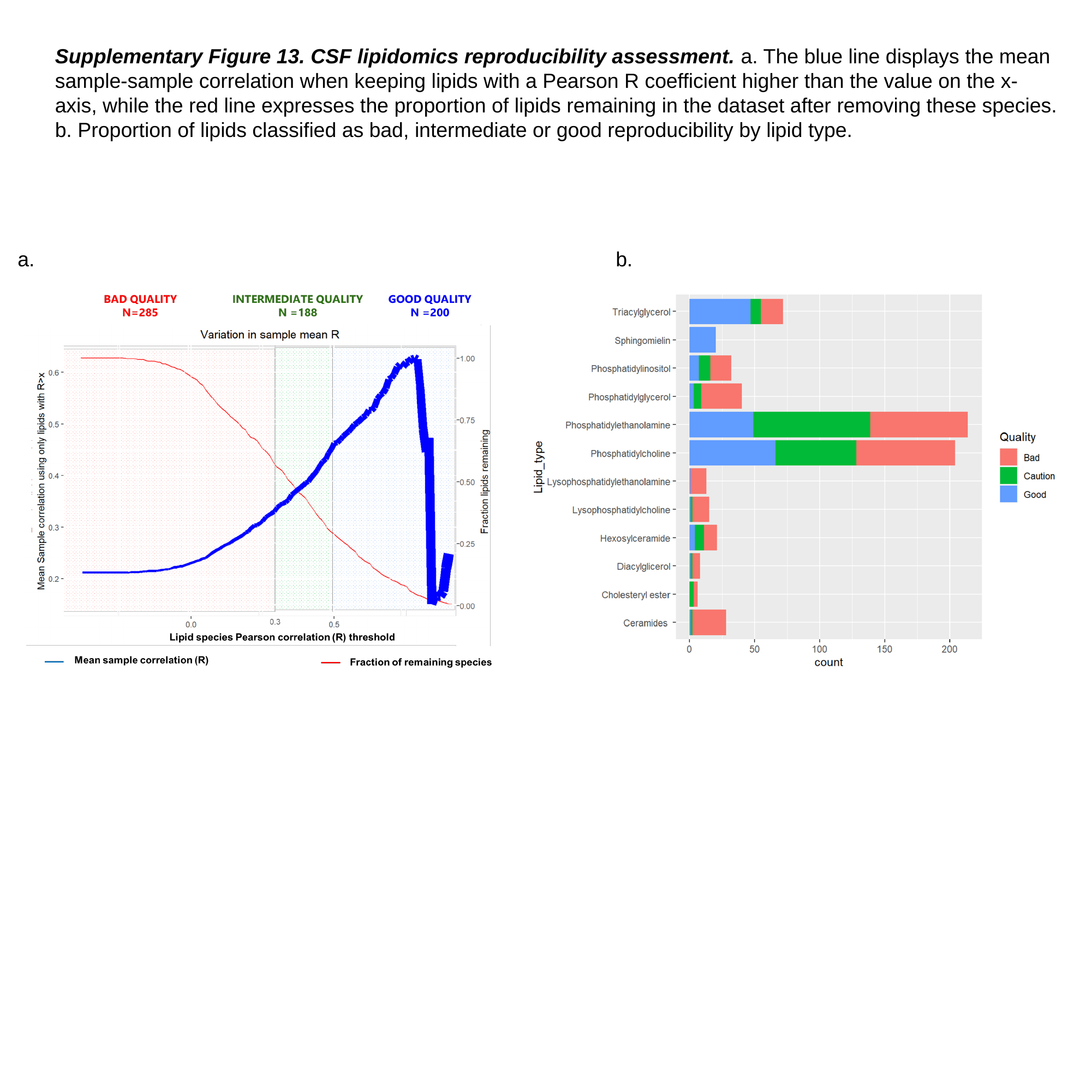

Supplementary Figure 13. CSF lipidomics reproducibility assessment. a. The blue line displays the mean sample-sample correlation when keeping lipids with a Pearson R coefficient higher than the value on the x-axis, while the red line expresses the proportion of lipids remaining in the dataset after removing these species. b. Proportion of lipids classified as bad, intermediate or good reproducibility by lipid type.
a.
b.
